# SARS-CoV-2 introductions to the island of Ireland

**DOI:** 10.1101/2023.05.11.23289783

**Authors:** Alan M. Rice, Evan P. Troendle, Stephen Bridgett, Behnam Firoozi Nejad, Jennifer M. McKinley, The COVID-19 Genomics UK consortium, National SARS-CoV-2 Surveillance & Whole Genome Sequencing (WGS) Programme, Declan T. Bradley, Derek Fairley, Connor G. G. Bamford, Timofey Skvortsov, David A. Simpson

## Abstract

Severe acute respiratory syndrome coronavirus 2 (SARS-CoV-2) has had an unprecedented impact on the people of Ireland as waves of infection spread across the island during the global coronavirus disease 2019 (COVID-19) pandemic. Viral whole-genome sequencing (WGS) has provided insights into SARS-CoV-2 molecular mechanisms of pathogenicity and evolution and contributed to the development of anti-virals and vaccines. High levels of WGS have enabled effective SARS-CoV-2 genomic surveillance on the island of Ireland, leading to the generation of a sizeable data set with potential to provide additional insights into viral epidemiology. Because Ireland is an island, accurate documentation of travel rates to and from other regions, both by air and by sea, are available. Furthermore, the two distinct political jurisdictions on the island allow comparison of the impact of varying public health responses on viral dynamics, including SARS-CoV-2 introduction events.

Using phylogenomic analysis incorporating sample collection date and location metadata, we describe multiple introduction and spreading events for all major viral lineages to the island of Ireland during the period studied (March 2020–June 2022). The majority of SARS-CoV-2 introductions originated from England, with frequent introductions from USA and northwestern Europe. The clusters of sequences predicted to derive from discrete introduction events (“introduction clusters”) vary greatly in size, with some involving only one or two cases and others comprising thousands of samples. When introduction cluster samples are mapped sequentially by collection date, they appear predominantly in previously affected or adjacent areas. This mirroring of the phylogenetic relationships by the geospatiotemporal propagation of SARS-CoV-2 validates our analytic approach. By downsampling, we estimate the power to detect introductions to Ireland as a function of sequencing levels. Per capita normalisation of both sequencing levels and detected introductions accounts for biases due to differing sequencing efforts and total populations. This approach showed similar rates of introductions for all major lineages into Northern Ireland (NI) and Republic of Ireland (RoI) with the exception of Delta, which was higher in NI which is likely attributable to higher travel per capita. However, there were similar rates of Delta infection within NI and RoI, suggesting that although travel restrictions will reduce risk of introducing novel variants to the region, they may not substantially decrease total incidence.

Our generalisable methodology to study introduction dynamics and optimal sequencing levels will assist public health authorities to select the most appropriate control measures and viral sequencing strategy.

## Introduction

Severe acute respiratory syndrome coronavirus 2 (SARS-CoV-2) is a non-segmented, single-stranded positive-sense RNA virus in the family *Coronaviridae*, which was first detected in Wuhan (武汉), Hubei (湖北) province, China (中华人民共和国) in December 2019, with an initial draft genome posted in early January 2020 (Andersen et al. 2020; Virological 2020). The virus is characterised by having an unusually large genome (~30 kbp) for an RNA virus, but is typical of coronaviruses (Brian and Baric 2005).

Despite the presence of an error-correcting RNA-dependent RNA replicase (Pachetti et al. 2020), which reduces the overall mutation rate (e.g., compared to human immunodeficiency virus or influenza viruses), SARS-CoV-2 still mutates with an estimated rate of about 1 x 10*^−^*^6^ to 2 x 10*^−^*^6^ mutations per nucleotide per replication cycle (Amicone et al. 2022; Gonźalez-Vázquez and Arenas 2023; Markov et al. 2023). RNA editing carried out by host proteins such as members of the ADAR gene family deaminating adenosine to inosine and APOBEC gene family deaminating cytosine to uracil result in pervasive A→G and C→U mutations (Di Giorgio et al. 2020; Simmonds 2020). This accumulation of viral mutations leads to an estimated substitution rate of about two novel single-nucleotide polymorphisms (SNPs) appearing each month (Duchene et al. 2020; Castillo-Morales et al. 2021; Markov et al. 2023). The continuous evolution of SARS-CoV-2 results in the sporadic emergence of new viral lineages, which can evade naturally acquired immunity and diminish the effectiveness of existing vaccines (Wang et al. 2022). New lineages are often characterised by many novel mutations that have accumulated rapidly prior to detection (Harvey et al. 2021) or, more rarely, by RNA recombination events (Li et al. 2020; Turakhia, Thornlow, A. Hinrichs, et al. 2022).

Whole-genome sequencing (WGS) facilitates the identification of viral variants, enabling viral transmission to be tracked in epidemics and pandemics (Baillie et al. 2012; Dudas et al. 2017; Grubaugh, Ladner, Kraemer, et al. 2017). Knowledge of circulating SARS-CoV-2 variants informs public health policies and responses (Talic et al. 2021). In response to the pandemic, the COVID-19 Genomics UK (COG-UK) consortium was established in April 2020 to provide high-throughput WGS of SARS-CoV-2 RNA extracted from positive cases in the nations of the United Kingdom (UK) (The COVID-19 Genomics UK (COG-UK) Consortium 2020). As a COG-UK member, we were responsible for sequencing the majority of Northern Irish SARS-CoV-2 samples (Fuchs et al. 2022) and also, in collaboration with members of the Sanger Institute, developed a dashboard for visualising dispersion of SARS-CoV-2 lineages across Northern Ireland (http://go.qub.ac.uk/covid19).

In the Republic of Ireland, SARS-CoV-2 WGS was predominantly performed by the National Virus Reference Laboratory in University College Dublin on behalf of the National SARS-CoV-2 Surveillance & WGS programme. The high per-capita SARS-CoV-2 WGS programmes within the island of Ireland provide an opportunity to study viral introductions into and spread between its two most populous urban centres (Belfast and Dublin) and the remainder of the island.

The island of Ireland (Irish: Éire, referred to hereafter as Ireland) is located on the periphery of northwestern Europe and offers a unique perspective to the study of viral transmission. Ireland has a significant amount of travel, not only with other European countries (O’Connor and Lyons 2003), but also with North America due to historical population migrations (Kenny 2014). Also, within Ireland there are two countries with a combined population of almost 7 million: Northern Ireland (NI) with a population of ~1.9 million (Northern Ireland Statistics and Research Agency (NISRA; Irish: Gńıomhaireacht Thuaisceart Éireann um Staitistićı agus Taighde), *Census 2021 main statistics for Northern Ireland (phase 1)*, https://www.nisra.gov.uk/publications/census-2021-main-statistics-for-northern-ireland-phase-1), which is one of the nations of the United Kingdom (UK), and Ireland (referred to hereafter as the Republic of Ireland (RoI)) with a population of ~5.1 million (Central Statistics Office (CSO, Irish: An Phŕıomh-Oifig Staidrimh), https://data.cso.ie/table/FP001). This geopolitical structure provides an opportunity to examine the effect of independent pandemic response measures on SARS-CoV-2 introductions and onward transmission over the course of the pandemic thus far on otherwise similar geographic regions. Note that the designation “Irish” is used to describe all samples collected in Ireland.

The transmission and severity of SARS-CoV-2 have been shown to be influenced by various human factors, including population density and regional deprivation. Studies have identified population density as a potential factor in the transmission of the virus, with variations observed across different regions of NI (McKinley et al. 2021). Moreover, the risk of dying due to SARS-CoV-2 infection has been found to be associated with regional deprivation (Public Health England 2020).

Research has demonstrated that deprivation is a primary risk factor for SARS-CoV-2 deaths among care home residents in England (Bach-Mortensen and Degli Esposti 2021). Furthermore, the mortality rate for deaths involving SARS-CoV-2 has also been shown to be significantly higher in the most deprived areas of England (Office for National Statistics 2020). Additionally, socioeconomic deprivation was linked to a higher rate of critical cases and mortality due to SARS-CoV-2 in Scotland (Lone et al. 2021). These findings suggest that regional deprivation is a key factor that must be considered when assessing the impact of the pandemic within Ireland.

In NI, there is already evidence linking SARS-CoV-2 incidence rate to deprivation data (McKinley et al. 2021). The identification of SARS-CoV-2 introductions and monitoring of viral spread allows us the opportunity to investigate their links with deprivation. Such an approach may help in identifying areas in Ireland that are at higher risk of transmission and mortality, and inform targeted public health interventions. By identifying areas that are most vulnerable to the virus, policymakers and healthcare professionals can develop targeted interventions and allocate resources more effectively, ultimately reducing pandemic harm.

Multifarious studies have characterised the initial introduction of SARS-CoV-2 to specific regions (Gonzalez-Reiche et al. 2020; Díez-Fuertes et al. 2020; Deng et al. 2020; Gámbaro et al. 2020; Bedford et al. 2020; Moreno et al. 2020; Maurano et al. 2020; Paiva et al. 2020; Komissarov et al. 2021; Muenchhoff et al. 2021; da Silva Filipe et al. 2021; Lobiuc et al. 2021; Murall et al. 2021; Juscamayta-Ĺopez et al. 2021; Githinji et al. 2021; du Plessis et al. 2021; Tordoff et al. 2021; Mallon et al. 2021; Lemieux et al. 2021; McBroome et al. 2022; Borges et al. 2022), or variants of SARS-CoV-2 to regions (Stefanelli et al. 2020; Tang et al. 2021; Toovey et al. 2021; Alpert et al. 2021; Elliott et al. 2022; Michaelsen et al. 2022; Fonager et al. 2022; Nasir et al. 2022; Bukhari et al. 2023). A previous phylogenetic analysis of 225 Irish genomes from two pandemic waves suggested that multiple introductions of SARS-CoV-2 to Ireland from outside had taken place (Mallon et al. 2021). However, the extent and originating locations of these and other introductions to Ireland remain to be elucidated.

Here, we leveraged a global SARS-CoV-2 phylogeny of more than seven million sequences to identify such introductory events. During six key time periods throughout the pandemic, we quantified introductions ascribed to the following major viral lineages as designated by the World Health Organization (WHO) and Pango nomenclature (O’Toole et al. 2022) as assigned by the pangolin tool (O’Toole et al. 2021): early initial variants, Pango B.1.177, Alpha (Pango B.1.1.7), Delta (Pango B.1.617.2/AY), as well as two major Omicron lineages (i.e., Pango B.1.1.529/BA.1 and Pango BA.2). The results yield insights into SARS-CoV-2 epidemiology within Ireland during each analysed time period, including SARS-CoV-2 importation frequencies as well as the subsequent geographic spread of infections. The workflow presented here is generalisable (i.e., it can be applied to any geographic region and virus of interest given sufficient sequencing data is available) and therefore can inform and improve global public health responses to both present and future viral and other pathogens.

## Materials and Methods

### Data

### SARS-CoV-2 phylogenetic tree

A global SARS-CoV-2 phylogeny (Lanfear 2020) dated 20^th^ May 2022 was downloaded from GISAID EpiCoV^™^ (Khare et al. (2021), https://www.gisaid.org) on 24^th^ May 2022. This phylogenetic tree is based on the masked alignment of 7,603,547 high quality SARS-CoV-2 genome sequences (Figure S1).

### Introductions SARS-CoV-2 sequence metadata

Metadata including the date and country of sample collection for all GISAID sequences was downloaded on 4^th^ June 2022. We have employed the GISAID nomenclature to attribute samples to countries.

#### SARS-CoV-2 confirmed cases

The number of daily confirmed SARS-CoV-2 cases for NI and RoI were downloaded from the Coronavirus (COVID-19) in the UK dashboard (https://coronavirus.data.gov.uk) and Ireland’s COVID-19 Data Hub (https://covid-19.geohive.ie) respectively.

#### Passengers arriving to Ireland by air

Our study analyzed aviation data pertaining to passengers traveling to Ireland between March 2020 and June 2022 as sourced from NISRA (https://www.nisra.gov.uk/statistics/tourism/northern-ireland-air-passenger-flow-statistics) for NI and CSO (https://data.cso.ie/table/TAM05) for RoI. While CSO data for RoI was directionally-resolved, allowing raw use of arriving passenger numbers, monthly arrival passenger populations for NI were estimated by halving the published passenger flow values (i.e., monthly arrivals = <u>^monthly passengers^</u>).

### Passengers arriving to Ireland by sea

To monitor the arrival of short sea ferry passengers traveling from Great Britain (GB) to the island of Ireland, we considered routes originating from Cairnryan (Scotland), Fishguard (Wales), Holyhead (Wales), Liverpool (England), or Pembroke (Wales) and terminating at Belfast (NI), Dublin (RoI), Larne (NI), or Rosslare (RoI). We also compiled data on international sea passenger routes to Ireland, which included routes departing from Bilbao (Spain), Cherbourg (France), and Rosscoff (France). These routes led to Cork, Dublin, and Rosslare in RoI. To obtain the sea passenger numbers for RoI, we used data from the Central Statistics Office (CSO). Specifically, we utilised the sum of the air and sea monthly directionally-resolved passenger data (https://data.cso.ie/table/ASM03) and then subtracted the corresponding figures for air travel (https://data.cso.ie/table/TAM05).

### Population density for government districts in Ireland

Population data within RoI and NI were obtained from CSO (https://data.cso.ie/table/FP001) and NISRA (https://www.nisra.gov.uk/publications/census-2021-main-statistics-for-northern-ireland-phase-1), respectively. To compute the population densities, the population within each government district was divided by the corresponding geographic area of the district, as sourced by Ordnance Survey of Northern Ireland (OSNI, https://www.opendatani.gov.uk/dataset/osni-open-data-largescale-boundaries-local-government-districts-20121) for NI and Ordnance Survey Ireland (OSI; Irish: Suirbhéireacht Ordanáis Éireann, https://data-osi.opendata.arcgis.com) for RoI. See Table S1 for the values used in the computation of population density.

### Deprivation for government districts in Ireland

The population data of subgroups in NI are primarily available at the level of Super Output Areas (SOAs). These areas were developed by the Northern Ireland Statistics and Research Agency (NISRA) with the aim of improving small-area statistics, and they were first produced for the 2011 census outputs (Northern Ireland Statistics and Research Agency 2013). Multiple deprivation measures in SOAs are presented as rankings, with SOAs in NI ranked from 1 (most deprived) to 890 (least deprived) (Northern Ireland Statistics and Research Agency 2017). Ranked-based data is deemed unsuitable for measurements, thus an indirect calculation of the level of deprivation is necessary using alternative data such as income level.

SOA data can be disaggregated into grid cells using the dasymetric technique (Firoozi Nejad 2016). As such, this method can be applied to disaggregate deprivation data (e.g., income level) from the SOA level into grid cells, generating grid-based deprivation data across NI. These outcomes can be subsequently merged with administrative areas to estimate the level of deprivation within those regions.

In NI, The proportion of the population residing in households with equivalised income below 60% of the NI median (Northern Ireland Statistics and Research Agency 2017) at the SOA level is utilised as ametric for measuring deprivation (%). For this analysis, WorldPop data for the entirety of NI in 2020 is also utilised. The dataset is obtainable at a granular level of grid cells (100m cell at the Earth’s equator) for both the UK and Ireland, with the units representing the number of individuals per cell (Bondarenko et al. 2020). To determine the number of individuals inhabiting each SOA (2020), population grid cells are utilised. Subsequently, the % of individuals living in deprivation is converted into the “number of deprived individuals in SOA”, and a straightforward algebraic expression is employed to devise a weighted overlay technique for disaggregating the number of deprived individuals into grid cells:

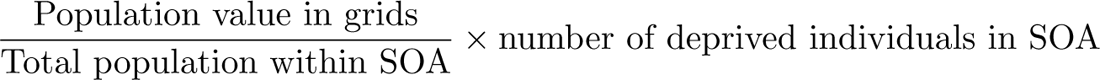

The results, which represent the number of deprived individuals in grid cells, are superimposed onto administrative boundaries to estimate the number of deprived individuals within each area. Subsequently, these figures are transformed into a percentage of deprived individuals within administrative areas, utilising total population data in each SOA.

Deprivation data (deprivation score) for the RoI is accessible at the administrative level of county councils (Teljeur et al. 2019). It is crucial to note that, in this study, the calculation of deprivation in NI and RoI is based on distinct methodologies, and therefore, direct comparisons between the two cannot be made.

### Country centroid positions

World country centroid positions were downloaded from https://github.com/gavinr/world-countries-centroids/releases/tag/v0.2.0. In addition to these, centroid positions were added individually for England (Lat: 52.561928 °N, Lon: 1.464854 °W), Scotland (Lat: 56.816738 °N, Lon: 4.183963 °W), Wales (Lat: 52.33022 °N, Lon: 3.766409 °W), and NI (Lat: 54.607577 °N, Lon: 6.693145 °W) from https://en.wikipedia.org/wiki/Centre_points_of_ the_United_Kingdom, as well as for Taiwan (https://en.wikipedia.org/wiki/Geographic_center_ of_Taiwan), Hong Kong (https://en.wikipedia.org/wiki/Module:Location_map/data/Hong_Kong), Ŕeunion (https://en.wikipedia.org/wiki/R%C3%A9union), Canary Islands (https://en.wikipedia.org/wiki/Module:Location_map/data/Spain_Canary_Islands), Palestine (https://en.wikipedia.org wiki/Module:Location_map/data/Palestinian_territories), Crimea (https://en.wikipedia.org/ wiki/Module:Location_map/data/Crimea), and Kosovo (https://en.wikipedia.org/wiki/Module: Location_map/data/Kosovo).

### Irish SARS-CoV-2 genomes and metadata for substitution rate estimation

The entire GISAID FASTA and metadata databases were downloaded on 18^th^ February 2023 to analyse population-wide SARS-CoV-2 genomics in Ireland. These data were then filtered to extract Irish samples (i.e., country=‘Ireland’ or country=‘NorthernIreland’) using augur filter v21.0.1 (Hadfield et al. 2018).

### Code availability

The analysis of data was conducted in Jupyter Notebook (https://jupyter.org); the analysis note-books used in this study can be accessed (https://github.com/QUB-Simpson-lab/paper-sars-cov-2-intros-to-ireland).

### Data availability

GISAID EPI ISL codes for all sequences analysed are provided (https://github.com/QUB-Simpson-lab/ paper-sars-cov-2-intros-to-ireland/analysis/datasets/GISAID/sequences.txt). A companion website (https://qub-simpson-lab.github.io/SARS-CoV-2-Ireland), which includes video files S13– S17 showcasing visualizations of geospatial introduction and spreading events of SARS-CoV-2, accompanies this publication. Our datasets associating population density and deprivation in Ireland to SARS-CoV-2 introductions and spread can are available (http://go.qub.ac.uk/gis-intros-to-ireland).

### Identifying and removing tree outliers

To identify samples that potentially have incorrect collection dates in their associated metadata or deviate substantially from their expected position in the tree under a molecular clock, the GISAID phylogenetic tree and sample collection dates were input to Chronumental v0.0.50 (Sanderson (2022), https://github.com/theosanderson/chronumental) to estimate a time-tree from the phylogeny. The difference between sample collection dates and the date predicted by Chronumental was calculated and a Z-score was calculated for each sample. Samples with a Z score greater than +3 or less than −3 were excluded from further analysis. These Z scores equate to roughly a ± 60 days difference in date. This step excluded 0.6% (46,278/7,603,547) of samples in the tree (Figure S2).

### Creating subtrees

GoTree v0.4.3 (Lemoine and Gascuel (2021), https://github.com/evolbioinfo/gotree) was used to prune the GISAID phylogenetic tree to subtrees used in subsequent analyses. Some NI samples were suspected to be erroneously labelled as being from NI. These “NI samples” with IDs other than ‘NIRE-’ were excluded when subtrees were created. Non-NIRE samples account for 5,427 NI samples.

### Reconstructing ancestral location of lineages

To infer ancestral locations of samples, a phylogenetic tree and the country of each sample in the tree was provided to PastML. Locations, at the level of countries, was inferred for each ancestral node of the phylogenetic tree (internal points between tips and the root of the tree that represent an ancestral state) using the DELTRAN (delayed transformation) method (Swofford and Maddison 1987) in PastML v1.9.34 (Ishikawa et al. (2019), https://github.com/evolbioinfo/pastml). Additionally, the --resolve polytomies PastML option was enabled.

### Clustering of sequences to identify independent introductions to Ireland

DendroPy v4.5.2 (Sukumaran and Holder (2010), https://dendropy.org) was used to traverse the output trees from PastML to identify Irish tips of the tree that were descended from ancestral nodes with a non-Irish country label assigned by PastML. We define all Irish tips linked to an importation event to comprise an introduction cluster.

The following periods were selected to span from prior to the first detected sequence of each major lineage in Ireland until they reached dominance.

### Period A - Initial introductions

To identify initial introductions to Ireland, the tree was pruned to include only sequences from the start of the pandemic up to the end of May 2020 and rooted using Wuhan-Hu-1 (GISAID: EPI ISL 402125) as an outgroup. Rooting a tree is the process of deciding which part of the tree is the oldest point and gives the direction of evolution moving from the root to the tips of the tree (Figure S3). An outgroup, a tip or lineage outside the group of tips of interest yet related to the the group, allows the root to be determined. Here, we use one of the earliest SARS-CoV-2 sequences as the outgroup to the rest of sequences in the tree. This tree included 103,316 tips of which 714 are from the RoI and 633 are from NI (Figure S4). Ancestral location of lineages was inferred by PastML as above. Location could not be resolved for 64 nodes (0.05%) and 2,053 new internal nodes were created by resolving polytomies.

### Period B - B.1.177

To identify Pango (O’Toole et al. 2022) lineage B.1.177 introductions to Ireland, the tree was pruned to include only B.1.177.* sequences (and renamed sublineages of B.1.177.*: AA.1, AA.2, AA.3, AA.4, AA.5, AA.6, AA.7, AA.8, Z.1, Y.1, W.1, W.2, W.3, W.4, V.1, V.2, U.1, U.2, U.3) from the first non-outlier occurrence of B.1.177 to the end of October 2020, and rooted using Wuhan-Hu-1 (GISAID: EPI ISL 402125) as an outgroup (Figure S5). This tree included 31,869 tips of which 215 are from RoI and 235 are from NI. Ancestral location of lineages was inferred by PastML as above. Location could not be resolved for 4 nodes (0.01%) and 364 new internal nodes were created by resolving polytomies.

### Period C - B.1.1.7 – Alpha

To identify Pango lineage B.1.1.7 introductions to Ireland, the tree was pruned to include only B.1.1.7 sequences (and renamed sublineages of B.1.1.7.*: Q.1, Q.2, Q.3, Q.4, Q.5, Q.6, Q.7, Q.8) from the first non-outlier occurrence of B.1.1.7 to the end of February 2021, and rooted using Wuhan-Hu-1 (GISAID: EPI ISL 402125) as an outgroup (Figure S6). This tree included 183,092 tips of which 2,850 are from RoI and 461 are from NI. Ancestral location of lineages was inferred by PastML as above. Location could not be resolved for 45 nodes (0.02%) and 2,181 new internal nodes were created by resolving polytomies.

### Period D - B.1.617.2/AY – Delta

To identify Delta introductions to Ireland, the tree was pruned to include only B.1.617.2/AY sequences from the first non-outlier occurrence to the end of July 2021, and rooted using Wuhan-Hu-1 (GISAID: EPI ISL 402125) as an outgroup (Figure S7). This tree included 512,420 tips of which 4,923 are from RoI and 1,613 are from NI. Ancestral location of lineages was inferred by PastML as above. Location could not be resolved for 287 nodes (0.05%) and 6,363 new internal nodes were created by resolving polytomies.

### Period E - B.1.1.529/BA.1 – Omicron

To identify Omicron introductions to Ireland, the tree was pruned to include only B.1.1.529/BA.1 sequences from the first non-outlier occurrence to the end of January 2022, and rooted using Wuhan-Hu-1 (GISAID: EPI ISL 402125) as an outgroup (Figure S8). This tree included 1,313,634 tips of which 7,888 are from the RoI and 3,615 are from NI. Ancestral location of lineages was inferred by PastML as above. Location could not be resolved for 497 nodes (0.03%) and 22,546 new internal nodes were created by resolving polytomies.

### Period F - BA.2 – Omicron

To identify BA.2 introductions to Ireland, the tree was pruned to include only B.2 sequences from the first non-outlier occurrence to the end of March 2022, and rooted using Wuhan-Hu-1 (GISAID: EPI ISL 402125) as an outgroup (Figure S9). This tree included 644,494 tips of which 1,235 are from RoI and 5,136 are from NI. Ancestral location of lineages was inferred by PastML as above. Location could not be resolved for 338 nodes (0.05%) and 12,722 new internal nodes were created by resolving polytomies.

### Downsampling tips from England

To determine the effect of the relative over-representation of English samples in the tree on the high level of English introductions identified to Ireland, we randomly downsampled tips from England in the tree. Between 10%-90% of English samples were randomly removed from the tree in 10% increments, with three replicates at each increment. Downstream analysis to identify introductions to Ireland was conducted as before.

### Downsampling tips from Ireland

To determine the effect of the level of sequencing in Ireland on the number and size of Irish introductions for the island as a whole and for each jurisdiction, we randomly downsampled tips from NI, RoI, and Ireland in the tree. Between 10%-90% of samples from Ireland were randomly removed from the tree in 10% increments, with one replicate at each increment. Downstream analysis to identify introductions to Ireland and clusters size was conducted as before.

### Estimation of total vs. identified introductions

A goal of this study was to evaluate the number of genetic sequences that would be required to identify all SARS-CoV-2 introductions to Ireland. To determine the additional number of introductions (*I*) that could be identified by analysing more sequences (*S*), we utilised ordinary least-squares (OLS) linear regression function scipy.stats.linregress from the Python library SciPy (Virtanen et al. 2020).

For each period, let S be the total number of available sequences and I be the corresponding number of introductions to Ireland identified. During downsampling, it is appropriate to select *N* (e.g., 10) evenly-spaced downsampled sequencing values *s_i_* such that

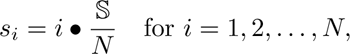

which yields an equivalent array

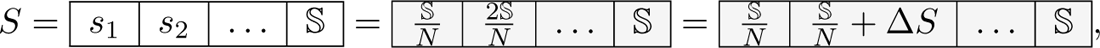

where 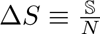

Through performing randomised downsampling of Irish tips in the phylogeny to each of the levels in array *S* and detecting introductions for each, a corresponding array for introductions, *I*, will be obtained:

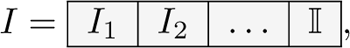

where *I*_1_ is the introductions predicted by downsampling to level *s*_1_, etc.

A numerical difference array (with size of *N −* 1) can be calculated from I:

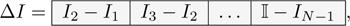

which enumerates the successively decreasing additional introductions identified upon increasing sequencing by Δ*S*.

We suppose that these diminishing returns of increased sequencing will continue to hold, such that at a sequencing saturation point S_all_, all predicted introductions, I_all_, would be identified and there would be limited additional value to increasing sequencing levels.

To estimate S_all_, the sequencing level required to predict all introductions, we perform OLS linear regression to assess the relationship:

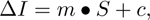

where *m* represents the slope of the linear regression line and *c* represents the intercept of the linear regression line. Note that as *S* has one element fewer than Δ*I*, we must prepend an element to Δ*I*, such that the first element contains the value *I*_1_, as the number of additional introductions identified from zero sequencing to the level of *s*_1_ is *I*_1_.

To apply the regression equation for extrapolation, values for *S* greater than the number of sequences obtained (S) may be input to compute new values for Δ*I*. Therefore, increased sequencing will provide additional introductions until Δ*I* = 0, where *S* = S_all_, which is the intersection of the regression line with the *S* axis: 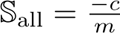

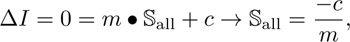

thereby creating an upper bound for the extrapolation of *S*.

To calculate I_all_, extrapolated *S* values (*S*_extrapolation_) are firstly generated in an array from S + Δ*S* to S_all_:

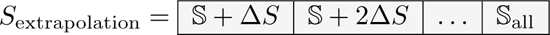

Inputting these (*S*_extrapolation_) values into the regression equation, we similarly obtain Δ*I*_extrapolation_:

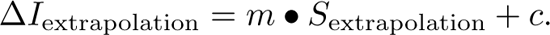

Each extrapolated Δ*I* value within Δ*I*_extrapolation_ is summed with I to predict I_all_, the total number of introductions to Ireland within the period.

Note that our analysis assumes that the number of additional introductions is directly proportional to the total number of sequences. However, this assumption may not hold true, and future research in this area will need to be undertaken to determine the ideal functional form with which to perform such extrapolations.

### Geospatial mapping

GeoPandas v0.11.1 (Jordahl et al. (2022), https://geopandas.org) was used to facilitate geospatial visualisations of SARS-CoV-2 introductions to Ireland. As source data, two geospatial vector data files (i.e., shapefiles) were downloaded from the GADM Database of Global Administrative Areas (https://gadm.org), which aims to provide high-quality maps of the administrative areas of all countries, at all recognised levels of sub-division to be utilised freely for non-commercial use. The shapefile gadm41 IRL 1.shp as found within the shapefile package downloadable from https://geodata.ucdavis.edu/gadm/gadm4.1/shp/gadm41_IRL_shp.zip was used to map the counties of RoI, while gadm41 GBR 2.shp as found within the shapefile package downloadable from https://geodata.ucdavis.edu/gadm/gadm4.1/shp/gadm41_GBR_shp.zip was used to map the local government districts (LGDs (2014), https://www.nisra.gov.uk/publications/nisra-geography-fact-sheet) of NI. The geographic resolution of RoI county and NI LGD is identical to the metadata fields accessible in GISAID or COG-UK metadata, allowing each sample to be associated to a distinct geographic region within Ireland. To create a map for the entire island of Ireland, NI was extracted from the shapefile containing the entirety of the UK and was merged with the shapefile containing RoI. Due to limited fine-detail information, sequenced samples were estimated to originate from the most populous settlement of each district as determined by Northern Ireland Statistics and Research Agency (NISRA; Irish: Gńıomhaireacht Thuaisceart Éireann um Staitistićı agus Taighde, https://www.nisra.gov.uk/publications/census-2021-main-statistics-for-northern-ireland-phase-1) for NI cities or Central Statistics Office (CSO; Irish: An Phŕıomh-Oifig Staidrimh, https://data.cso.ie/table/FP001) for RoI cities. Latitudes and longitudes of these representative cities were obtained from Microsoft Bing Maps (https://www.bing.com/maps). To annotate the maps, Troendle–Simpson–Skvortsov 2-letter abbreviation codes were devised for each local government district as indicated in Table S1.

### Spatial statistical analysis of Geographic Information System (GIS) data

Spatial statistical analyses of Geographic Information System (GIS) data were automated by the Spatial Statistics toolbox in the ArcGIS Pro v3.0.2 (Build 3.0.2.36056) software developed by Esri (https://www.esri.com/en-us/arcgis/products/arcgis-pro/overview). Geospatial data was input to the Spatial Statistics toolbox in ArcGIS Pro in the form of shapefiles, geodatabases, or feature classes, containing both the attribute and location data of the features being analysed.

The Ordinary Least Squares (OLS) functionalities used within the ArcGIS Pro Spatial Statistics toolbox provide the results of bivariate regression of two independent variables: population density, and deprivation in correlation with the dependent variable: the proportion of Irish tips sequenced in the region linked to introduction and spreading events within the periods studied. The specific statistical measures revealed by the OLS functionalities are p-values, robust p-values, adjusted R^2^s, variance inflation factors (VIFs), and Koenker (BP) statistic (i.e., Koenker’s studentised Bruesch-Pagan statistic) probabilities.

### SARS-CoV-2 genomic trends in Ireland

We separately aligned each Irish genomic sequence from GISAID with Pango lineages corresponding to those of the six periods studied (*N* =140,121) to the Wuhan-Hu-1 reference genome (GISAID: EPI ISL 402125) using MAFFT v7.453 (Katoh et al. 2002). The number of substitutions in each Irish genome compared to the Wuhan-Hu-1 reference genome was determined by calculating the Hamming distance (Hamming 1950), which tallies the minimum number of substitutions required to change the reference sequence into a subject sequence of equal length, while excluding the contributions of insertions, deletions, and Ns in the subject sequence.

To analyse substitution rates per period, we conducted ordinary least squares (OLS) linear regression using the LinearRegression model from scikit-learn v1.1.3 (Pedregosa et al. (2011), https://scikit-learn.org). The data in Table S8 includes error estimates and p-values from regressions that were nearly identical, but conducted using the OLS functionality in statsmodels v0.13.5 (Seabold and Perktold (2010), https://www.statsmodels.org).

## Results

Confirmed SARS-CoV-2 cases followed similar trends in NI and RoI, with some differences including more cases in NI in autumn 2021 and Feb 2022 and more in RoI in Jan 2022 (Figure 1A), which may reflect disparities in viral epidemiology and/or pandemic response measures between the two jurisdictions. The number of samples sequenced per month was mirrored between NI and RoI, with the per capita rate being higher in NI (Figure 1B). Successive waves of dominant SARS-CoV-2 Pango (O’Toole et al. 2022) lineages were very similar, with a few other lineages persisting in RoI after the introduction of Alpha and a delayed rise in Omicron BA.2 in NI (Figure 1C). The introduction of these lineages to the island of Ireland will be our focus here.

**Figure 1.**
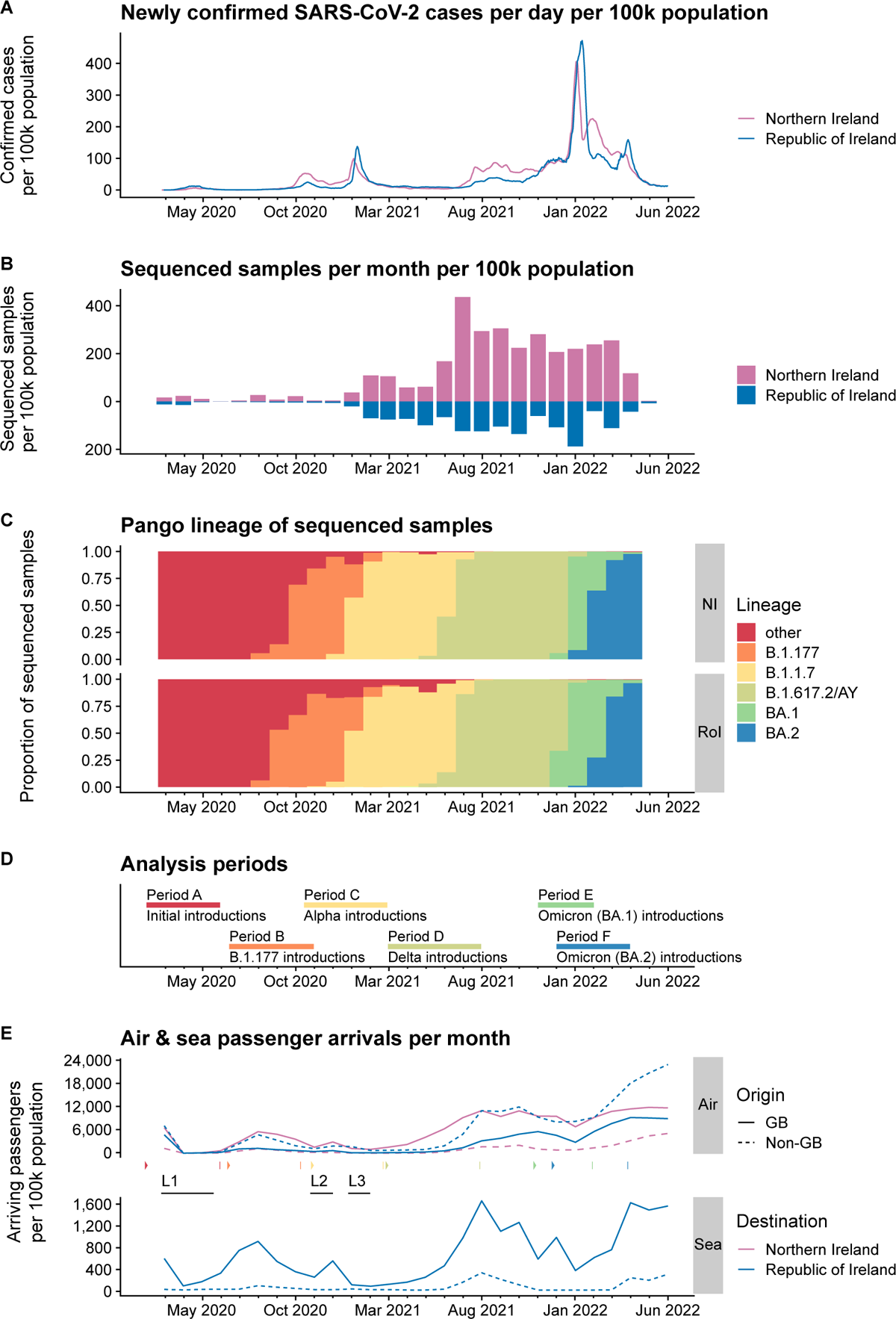
SARS-CoV-2 timeline in Northern Ireland and Republic of Ireland. **A**, Seven day rolling daily mean of confirmed SARS-CoV-2 cases per 100,000 population. Data from Coronavirus (COVID-19) in the UK dashboard and Ireland’s COVID-19 Data Hub. **B**, Monthly histogram by collection date of SARS-CoV-2 genome sequences deposited to GISAID for Northern Ireland (NI) and Republic of Ireland (RoI). **C**, Pango lineage of genome sequences deposited in GISAID for NI and RoI. Major lineages highlighted, all other lineages grouped under ‘other’. **D**, Analysis periods used in this study to identify SARS-CoV-2 introductions to the island of Ireland. **E**, Maritime and aviation passenger arrival volumes to NI and RoI per month. Maritime data for passenger arrivals into NI for the period studied is not presently available. Lockdowns in Ireland are represented by three periods (L1-3) that occurred from late March to mid May 2020, late October to December 2020, and late December 2020 to February 2021, respectively. Although there were slight variations in the particular measures implemented between NI and RoI, these dates were selected to demonstrate a general agreement on lockdown periods between the two regions. The starts (▷) and the ends (*|*) of the periods studied are indicated.

To estimate the number of SARS-CoV-2 introductions to the island of Ireland, we used a phylogenetic tree from May 2022 available from GISAID EpiCoV^™^ (Lanfear 2020). This tree was based on the masked alignment of 7,603,547 high quality SARS-CoV-2 genome sequences. To identify sequenced samples that have incorrect metadata collection dates or deviate substantially from their expected position in the tree under a molecular clock, a time-tree was estimated from the phylogeny. Outliers, equating to roughly a 60 days difference between metadata date and predicted date, were excluded from further analysis (Figure S2). This step excluded 0.6% (46,278/7,603,547) of sequences. A number of analysis periods, A to F, were chosen as critical periods during the SARS-CoV-2 pandemic when important viral importations occurred (Figure 1D).

Due to inconsistencies between country and fine-grain location metadata, some NI samples were suspected to be erroneously labelled as being from NI. This could overestimate the number of identified introductions. Samples with an NIRE prefix ID (e.g. NIRE-000107) should all be collected and sequenced in NI and correctly attributed a country location of NI. Samples with NIRE prefix IDs comprise 84.3% of NI samples in the phylogenetic tree, while RAND IDs comprise 8.2%, NORT IDs 2.7%, and 15 other prefixes total 4.8% of NI samples. Therefore, identified NI introductions were filtered to consider introductions that consisted of NIRE IDs only. This excluded 5,427 NI samples.

### SARS-CoV-2 introductions from Europe and USA seeded early pandemic

Despite the relatively low numbers of sequenced samples in both RoI and NI from 2020 (Figure 1B), some insights can be gleaned about the nature of these early importations. The first analysis period (Period A) is the beginning of the pandemic and aims to quantify the initial introductions of SARS-CoV-2 tothe island of Ireland (Figure 1C&D). The tree was pruned to include only sequences from the start of the pandemic up to the end of May 2020. This tree included 103,316 tips of which 714 are from RoI and 633 are from NI. A country was estimated for each ancestral lineage of the phylogenetic tree using the DELTRAN (delayed transformation) method. SARS-CoV-2 introductions to Ireland could then be inferred by Irish sequences that are descendants of a non-Irish ancestral node in the tree.

This method estimates that at least 170 independent introductions of SARS-CoV-2 took place in the time period up to the end of May 2020, 103 to RoI and 67 to NI, with three intra-Irish introductions, two from RoI to NI and one from NI to RoI. The majority of introductions originated from Europe, with 59.5% of introductions from England, 10.1% from the United States of America, and 4.8% from Scotland (Figure 2 Period A left, Table S2). There was a significant positive correlation between the number of introductions from a country and the number of tips in the tree from a country (*ρ* = 0.75, *P* = 0.002, Spearman’s rank correlation; Figure 2 Period A centre). There was no correlation between the number of introductions from a country and its distance from Ireland (*ρ* = −0.47, *P* = 0.45, Spearman’s rank correlation; Figure 2 Period A right).

**Figure 2.**
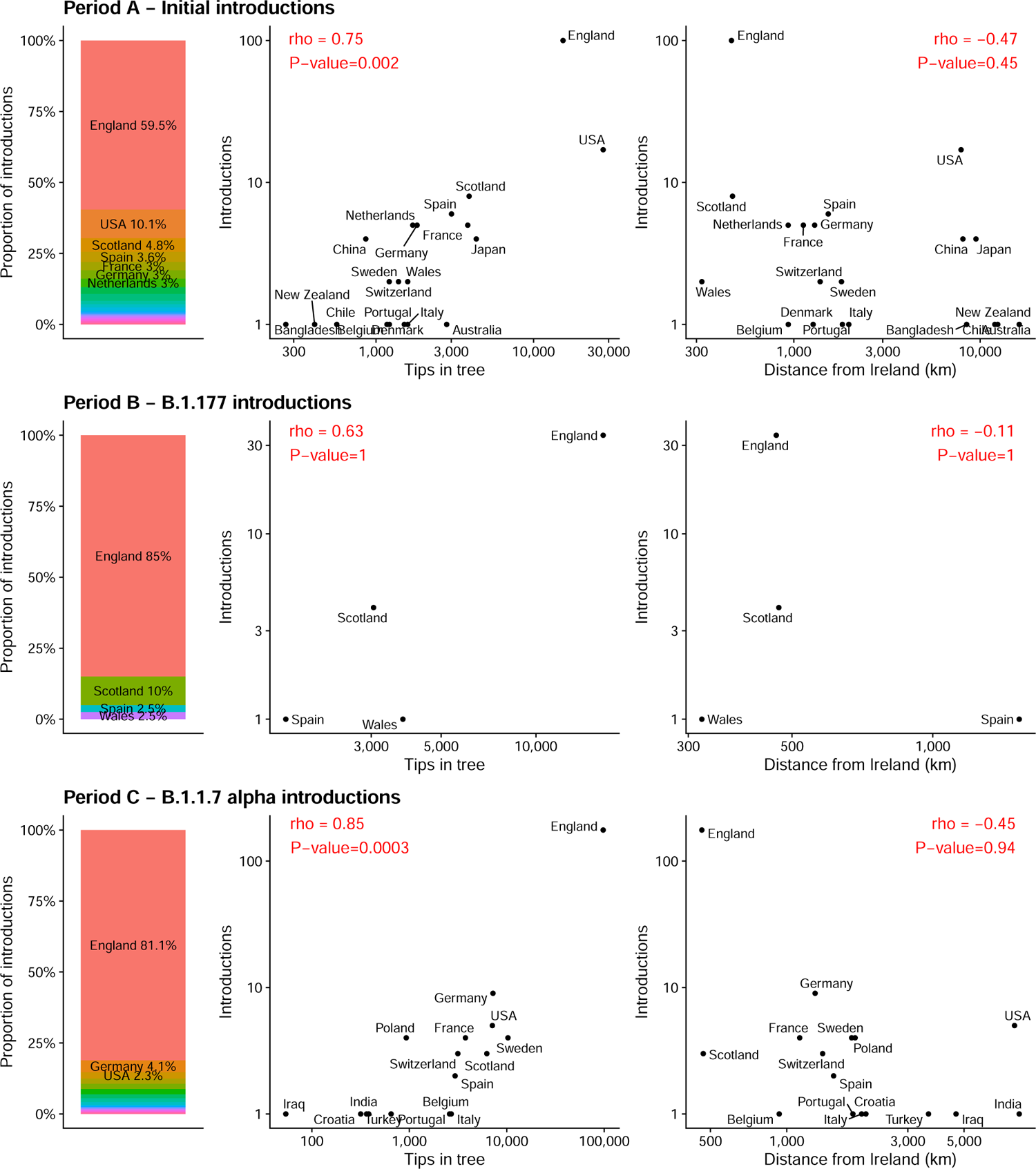
SARS-CoV-2 introductions for Periods A, B, and C. Left for each period, the proportion of importations from each country. Centre, the number of introductions and the number of tips per country in the tree. Right, the number of introductions per country and the distance in kilometres between Ireland and each respective country. Note the log_10_ scale for both the *x* and *y* axis. P-values for Spearman correlation tests are Bonferroni-corrected for multiple testing.

### B.1.177 and Alpha introductions originated predominantly from England

The second analysis period (Period B) focuses on the emergence of Pango lineage B.1.177 over summer 2020 until the end of October 2020 by which time B.1.177 had become dominant in Ireland (Figure 1C&D). The phylogenetic tree was pruned to contain B.1.177 sequences from this time and included 31,869 tips, of which 215 were from RoI and 235 were from NI. There were 41 introductions identified, of which 23 were into NI and 18 into RoI (Table S3). Almost all introductions originated from Great Britain, with 85% of introductions from England (Figure 2 Period B left). With only four unambiguous countries (England, Scotland, Wales, & Spain) for all importations in this period (Table S4), no meaningful correlation with number of tips in the phylogenetic tree and distance from Ireland could be performed (Figure 2 Period B centre and right).

Period C focuses on introductions of B.1.1.7 Alpha lineage until the end of February 2021 (Figure 1C&D). The phylogenetic tree was pruned to 183,092 tips of which 2,850 were from RoI and 461 were from NI. There were 173 introductions into the RoI from outside Ireland, and 45 into NI (Table S4). Additionally, there were 25 introductions between RoI and NI (14 from NI and 11 from RoI). The majority of introductions originated from England (81.1%), followed by Germany (4.1%), and USA (2.3%; Figure 2 Period C left). While there was no correlation between the number of introductions and the distance from Ireland, there was a significant positive correlation between the number of introductions from a country and the number of tips in the tree from a country (*ρ* = 0.85, *P* = 0.0003, Spearman’s rank correlation; Figure 2 Period C right and centre, respectively).

### Delta lineage imported from England, Europe, and India

#### Period D focuses on introductions of Delta lineages (Pango lineages B.1.617.2/AY) until the end of July 2021 (Figure 1C&D)

The phylogenetic tree was pruned to 512,420 tips of which 4,923 are from RoI and 1,613 are from NI. There were 454 introductions into RoI from outside Ireland, 273 into NI, and one introduction to either RoI/NI (Table S5). Additionally, there were 73 introductions between RoI and NI (31 from NI and 42 from RoI). The majority of introductions originated from England (66.1%), followed by Scotland (6.1%), India (5.3%), and Spain (3.4%; Figure 3 Period D left). Interestingly, these are not balanced between NI and RoI. There were 228 introductions from England to NI (83.5% of NI introductions) yet only 224 introductions to RoI (49.3% of RoI introductions). Conversely, there were 33 introductions from India to RoI (7.2% of RoI introductions), and 23 from Spain (5.1%) yet only 5 introductions from India to NI (1.8% of NI introductions) and 1 from Spain (0.4%).

**Figure 3.**
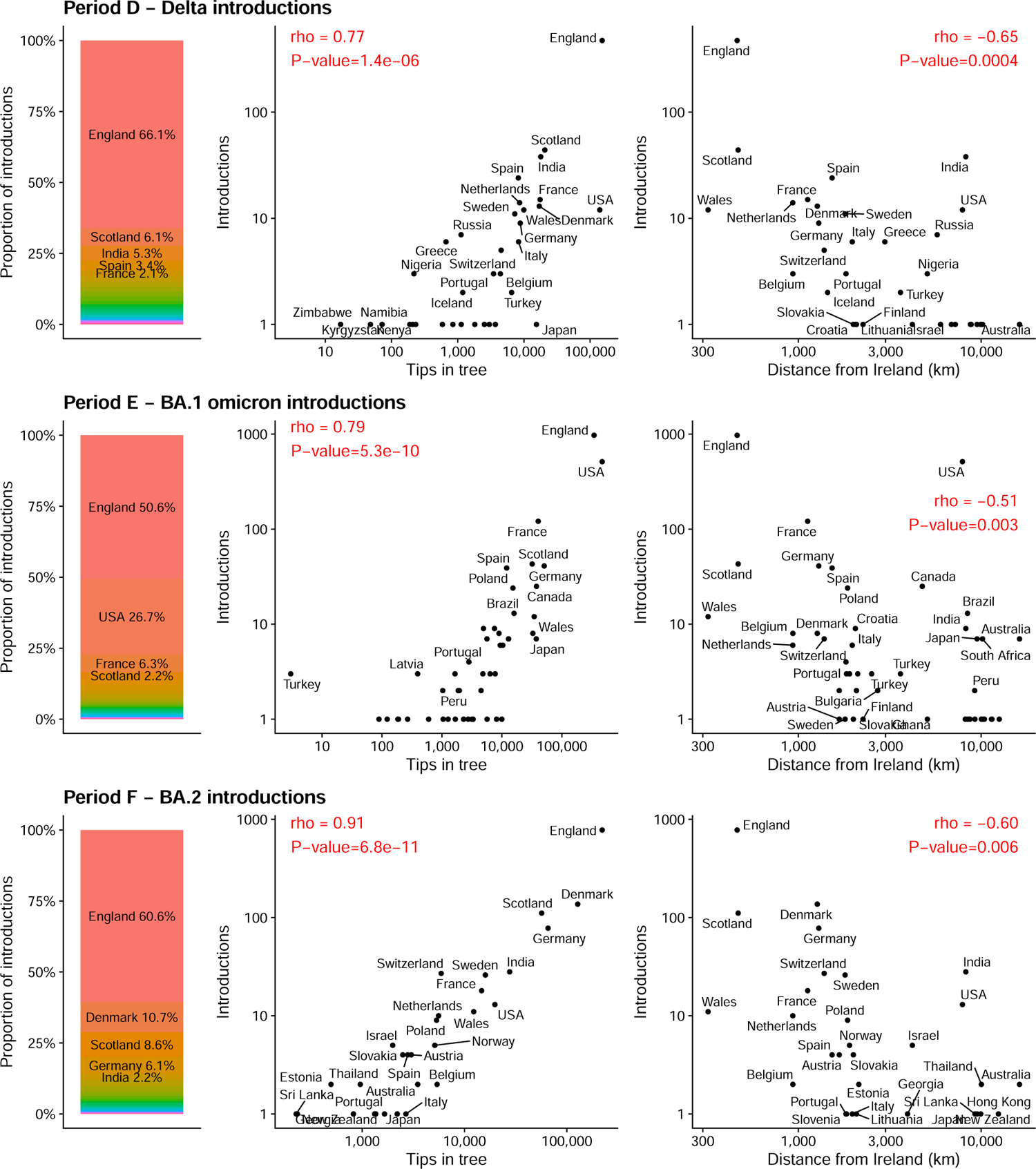
SARS-CoV-2 introductions for Periods D, E, and F. Left for each period, the proportion of importations from each country. Centre, the number of introductions and the number of tips per country in the tree. Right, the number of introductions per country and the distance in kilometres between Ireland and each respective country. Note the log_10_ scale for both the *x* and *y* axis. P-values for Spearman correlation tests are Bonferroni-corrected for multiple testing.

For Period D, we found a significant positive correlation between the number of introductions from acountry and the number of tips in the tree from that country (*ρ* = 0.77, *P* = 1.4 10*^−^*^6^, Spearman’s rank correlation; Figure 3 Period D centre). There was also a significant correlation between the number of introductions from a country and the distance of each country from Ireland (*ρ* = 0.65, *P* = 0.0004, Spearman’s rank correlation; Figure 3 Period D right).

### England and USA account for majority of BA.1 introductions

#### Period E focuses on introductions of the Omicron (BA.1) lineage (Figure 1C&D)

The phylogenetic tree was pruned to include 1,313,634 tips of which 7,888 were from RoI and 3,615 were from NI. There were 1,937 introductions to Ireland (1,329 introductions into RoI and 608 into NI) and 69 introductions between RoI and NI (31 from NI and 38 from RoI; Table S5). About half of introductions were from England (50.8%), followed by USA (26.7%), and France (6.3%; Figure 3E left). We found a significant positive correlation between the number of introductions from a country and the number of tips in the tree from that country for Period E (*ρ* = 0.79, *P* = 5.3 10*^−^*^10^, Spearman’s rank correlation; Figure 3 Period E centre). There was also a significant correlation between the number of introductions from a country and the distance of each country from Ireland (*ρ* = −0.51, *P* = 0.003, Spearman’s rank correlation; Figure 3 Period E right).

### BA.2 introductions predominantly from Northern Europe

#### Period F focuses on introductions of the Omicron (BA.2) lineage (Figure 1C&D)

The phylogenetic tree was pruned to include 644,494 tips of which 1,235 were from the RoI and 5,136 were from NI. There were 1,309 introductions to Ireland (428 introductions into the RoI and 881 into NI) and 34 introductions between RoI and NI (29 from NI and 5 from the RoI; Table S6). England (60.6%), Denmark (10.7%), Scotland (8.6%), and Germany (6.1%) were the most frequently observed origin countries for importations (Figure 3 Period F left).

For Period F, we found a significant positive correlation between the number of introductions from acountry and the number of tips in the tree from that country (*ρ* = 0.91, *P* = 6.8 10*^−^*^11^, Spearman’s rank correlation; Figure 3 Period F centre). There was also a significant correlation between the number of introductions from a country and the distance of each country from Ireland (*ρ* = −0.60, *P* = 0.006, Spearman’s rank correlation; Figure 3 Period F right).

### Clusters comprising multiple Irish sequences vary greatly in size

A cluster of multiple Irish sequences that arise from a single introduction event can be informative about virus spreading and epidemiology when combined with geographic or other information. However, 63.2% (2,782/4,403) of introductions across all six periods resulted in detection of only a single Irish descendant (Figure 4), with the percentage at specific periods varying from 38.1% (Period C) to 68.9% (Period E) between periods studied. The largest cluster in each period ranged from 124 sequences in Period B to 2,228 sequences in Period F. The large number of clusters having few samples implies that despite the globally high per capita sequencing rate in Ireland, the level of WGS sequencing may be insufficient to detect most cases spreading from each introduction and the number of importations will have been underestimated. Any conclusions regarding geographic spread must be tempered by this limitation and also take into account the variable proportion of positive cases sequenced as a result of changing case numbers and sequencing effort.

**Figure 4.**
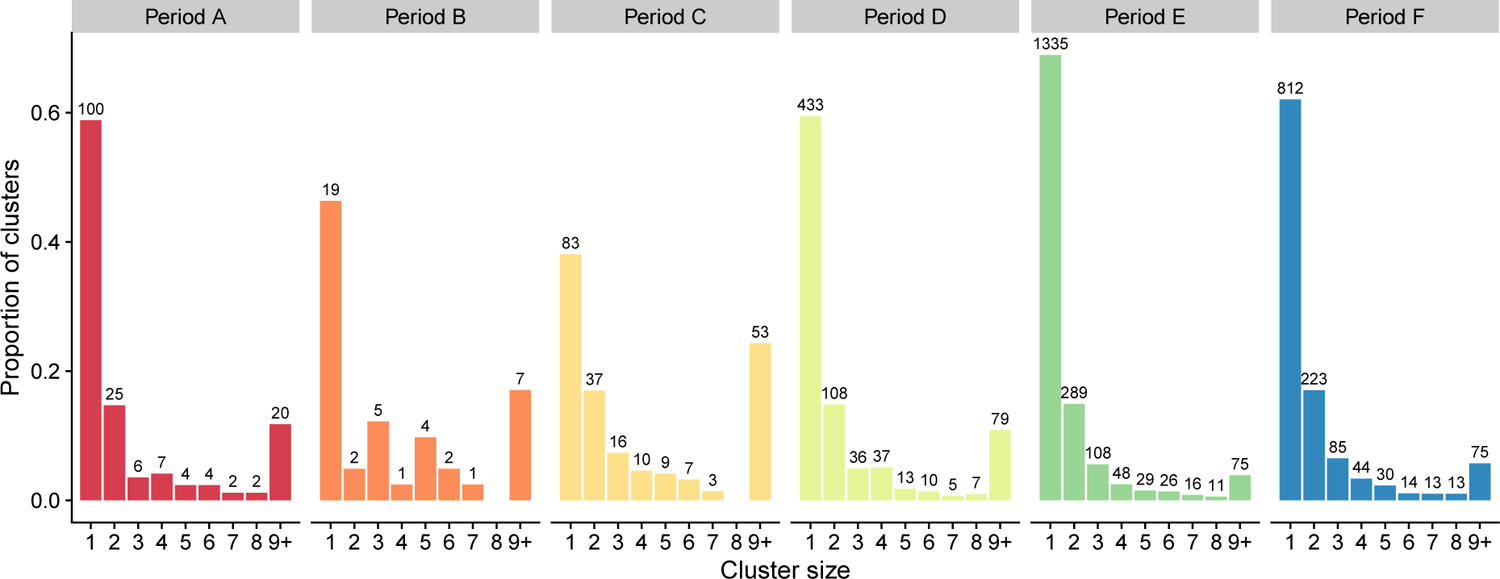
Irish samples per introduction. Number of Irish samples per introduction grouped by cluster size 1-9+ for Periods A–F. Bar height shows the proportion of total clusters for each cluster size within a period.

### Introductions from England exceed predictions based on sample numbers in the global phylogeny

Countries with an open, high-throughput variant surveillance program will feature more frequently in the phylogenetic tree than other countries that have made few sequences publicly available. There is astrong relationship between the number of tips in the tree from a specific country and the number of introductions from the same country (Figures 2 and 3). England is the most frequent origin country of importations to Ireland for all periods analysed. However, these high levels of importations observed could be due to England being a global leader in the SARS-CoV-2 sequencing effort by making many genome sequences available, rather than due to factors like the country being geographically close to Ireland with high connectivity.

To investigate the effect of sampling bias on the number of Irish importations attributed to England, we randomly downsampled tips from England in the tree to levels retaining between 90% and 10% of the original number and then counted importations to Ireland as before. If the proportion of importations from England to Ireland was due solely to the amount of English samples in the tree, we could expect a linear relationship when downsampling, i.e., reducing the number of English samples in the tree by 50% would reduce the proportion of introductions attributed to England by 50%. Instead, as samples are removed, we observe that the proportion of introductions from England remains higher than expected (Figure 5A). This is significant for all six periods when compared to the linear expectation (*P* between 8.9 x10*^−^*^8^ and 3.6 x10*^−^*^5^ for each period; paired Wilcoxon signed-rank test). Additionally, the proportion of introductions from England to Ireland is always greater than the proportion of English samples in the tree for all downsampled replicates (Figure 5B).

**Figure 5.**
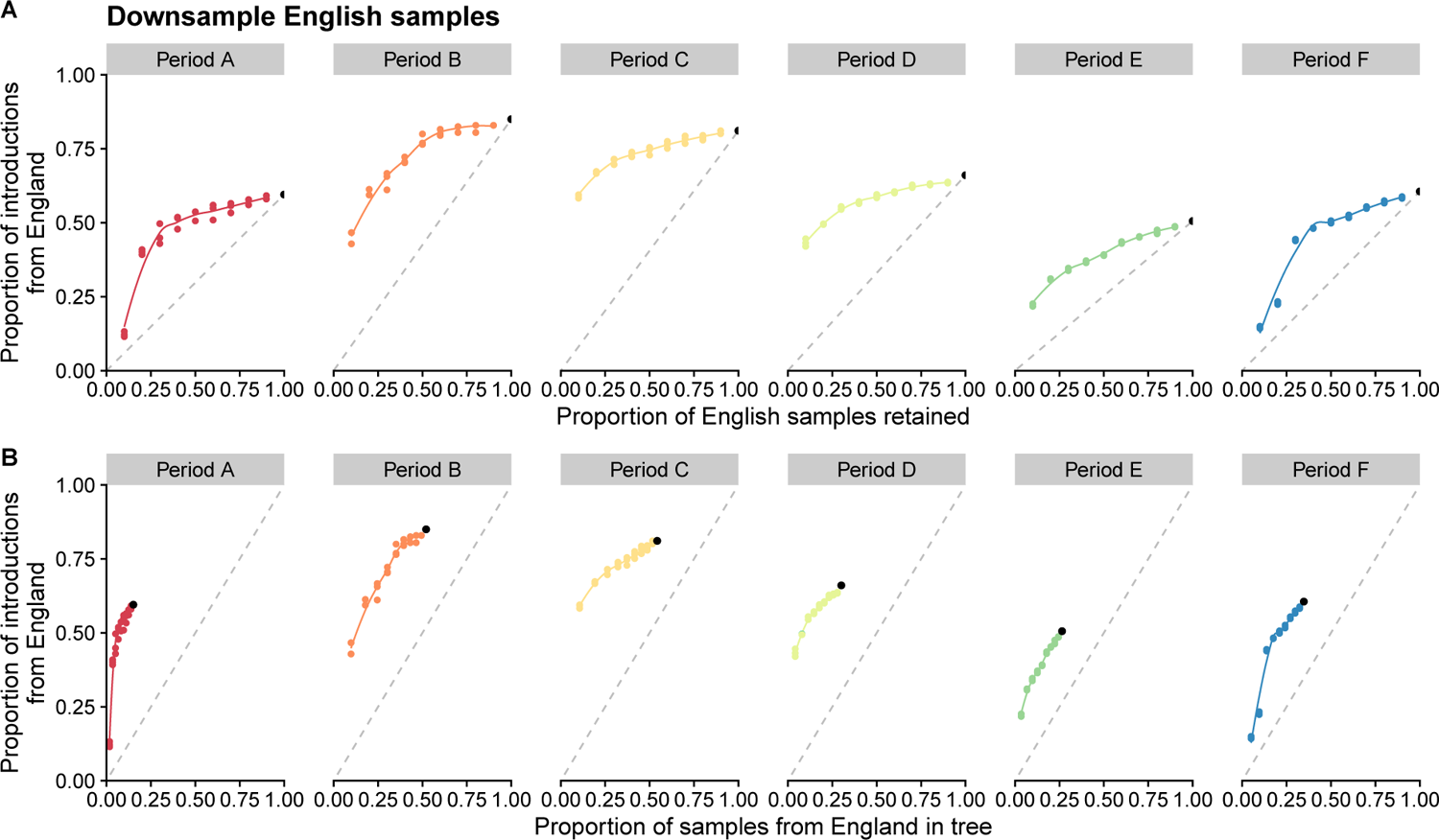
Effect of downsampling English samples. Three replicates for each downsampled proportion. The black point is the number of observed introductions for each period using all data. Dashed grey lines represents a linear relationship between zero and observed introductions. The smoothed lines are fit to the downsampled data points using local polynomial regression fitting (loess). **A**, Proportion of introductions to Ireland observed from England when English samples are randomly downsampled by 10% to 90%. **B**, Proportion of introductions to Ireland observed from England when English samples are randomly downsampled and the proportion of samples in the phylogenetic tree that are from England. Three replicates for each downsampled proportion. Dashed grey line represents a linear relationship between 0% and 100%.

### Optimal Irish sequencing rates vary between periods

Different levels of sequencing impact the ability to detect introduction events throughout the pandemic. Increased SARS-CoV-2 sequencing in both RoI and NI in 2021 (Figure 1B) yielded more rapid and informed surveillance of variants.

Sequencing at a very low level will miss whole clusters resulting from importations and so when sequencing is low, the number of importations will be underestimated. At the other end of the spectrum, sequencing every SARS-CoV-2 case will detect all members belonging to each cluster, resulting in the best possible estimate of the number of importations. However, as the level of sequencing increases, the returns are diminishing because different members of the same cluster would be sampled, in which case the number of importations remains much the same.

To determine the effect of different levels of sequencing on the number of Irish importations detected, we randomly downsampled Irish sequences in the tree, similar to above, retaining between 90% and 10% and then counted importations to Ireland as before. For Periods A–C, removing 90% of Irish samples, reduces the number of introductions detected to between 31% and 44% of the introductions observed (Figure 6A). This is greater than the 10% expectation if introductions decreased linearly. For Periods D–F, a more steep decline is observed as samples are removed and when only 10% remain, we detect between 22% to 27% of the original observed number of introductions. When average cluster size is considered, a steeper decline is instead observed for Periods A–C compared to D–F (Figure 6B). Therefore, we hypothesise that if sampling was increased in Ireland beyond the level conducted for Periods A–C, few additional introductions would be detected, but instead most new samples would be added to existing introduction clusters and increase cluster size. For Periods D–F, an increasing in sampling would continue to detect many new introductions and increase cluster size of existing introductions.

**Figure 6.**
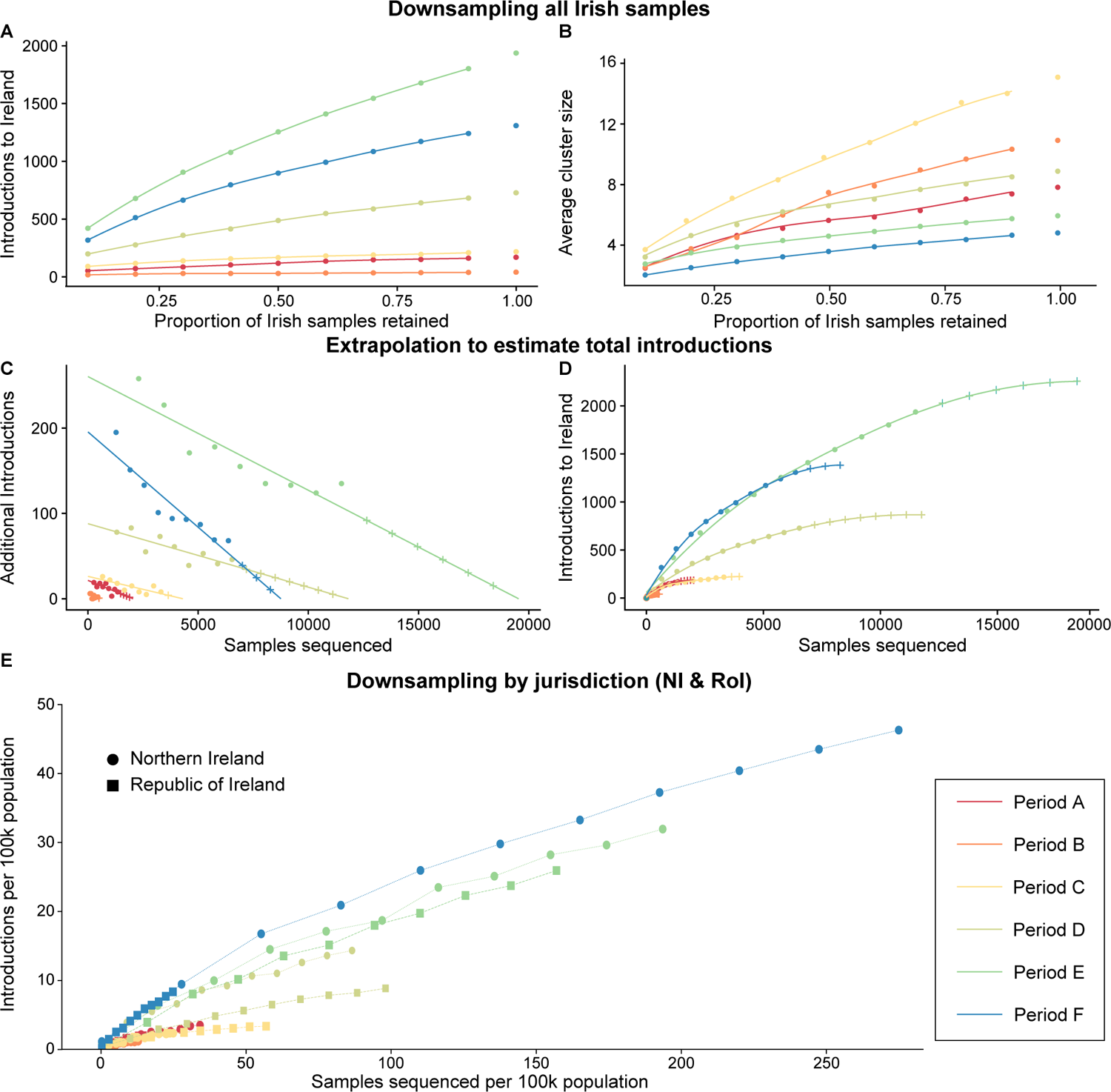
Effect of downsampling Irish samples. (A & B), corresponding extrapolation to estimate total introductions to Ireland (C & D), and population normalised introduction and sequencing rates for NI and RoI (E). A, Number of introductions to Ireland observed when Irish samples are randomly downsampled by 10% to 90%. One replicate for each downsampled proportion. B, Average cluster size of Irish introductions when Irish samples are randomly downsampled by 10% to 90%. C, The numerical derivatives of the downsampled data (A) were assessed and fit to a straight line to approximate the rate additional sequencing will have on finding additional introduction clusters. D, The data is extrapolated from C, revealing predicted sequencing saturation levels, where additional sequencing is unlikely to detect novel introduction events. E, Comparison of the number of introductions to NI and RoI for each period studied. Downsampling was performed independently for samples from each jurisdiction. To account for the difference in total populations, both the sets of downsampled sequences and identified introductions were normalised per 100k population.

**Figure 7.**
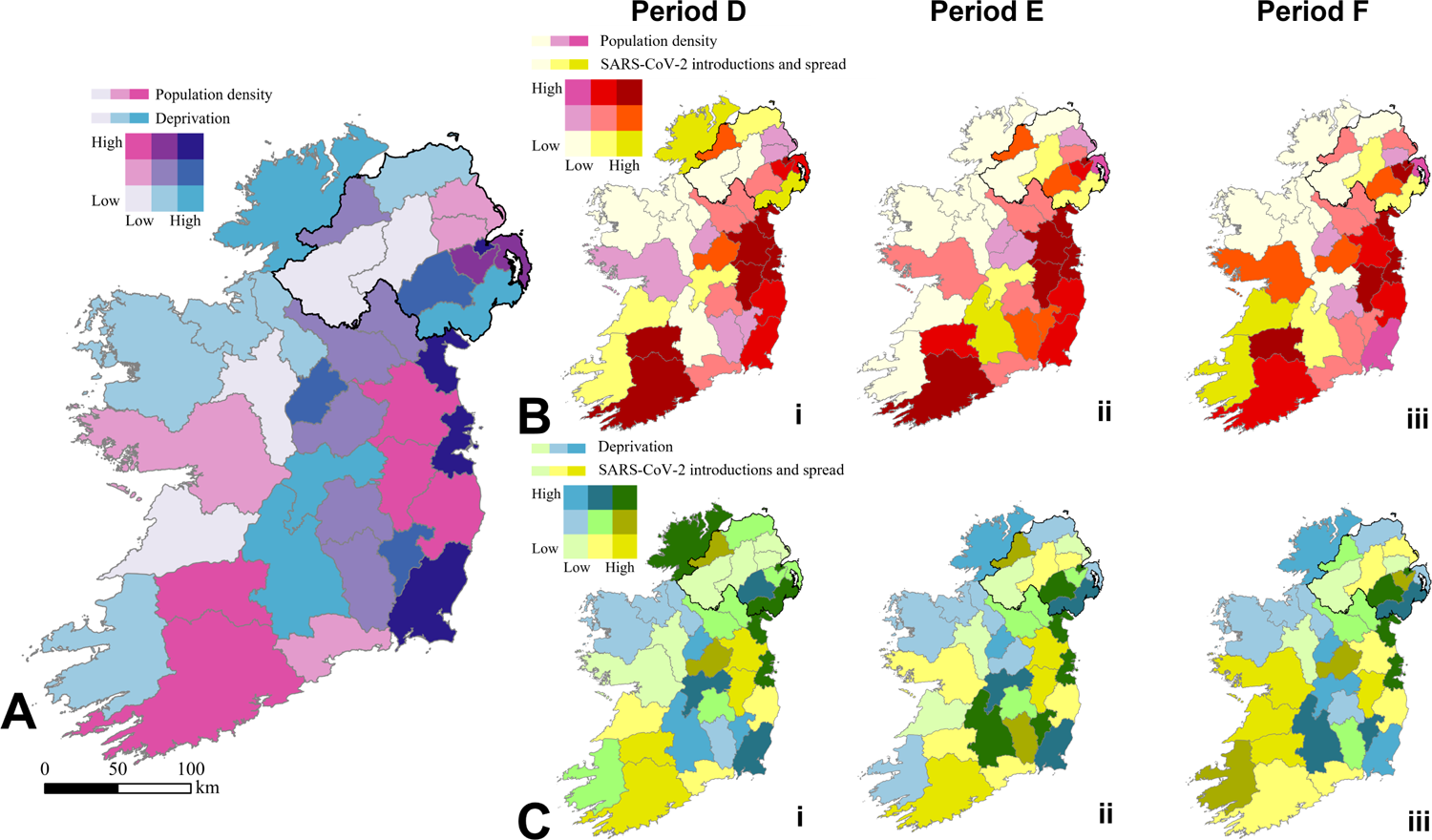
Bivariate sequential choropleth maps (9-class) depicting relationships between population density, deprivation, and SARS-CoV-2 introductions and spread for all local government districts in Ireland. A, The association between population density (cyan palette) and deprivation (magenta palette) in Ireland. It is assumed that this relationship remains constant throughout the periods studied. B, The correlations between population density (cyan palette) and the proportion of samples linked to the introduction and spread of SARS-CoV-2 (dark yellow palette). The three maps labeled i–iii correspond to periods D–F, respectively. C, The correlations between deprivation (magenta palette) and the proportion of samples linked to the introduction and spread of SARS-CoV-2 (dark yellow palette). The three maps labeled i–iii correspond to periods D–F, respectively.

It is a challenge for public health officials to assess the effectiveness of a given level of sequencing. To assist in this evaluation, we have developed a methodological framework involving the extrapolation of downsampled data to determine the proportion of total predicted introductions that were detected using the available sequencing. The extrapolation results (see Table 1 and Figure 6C&D) also reveal the estimated number of sequences required to identify all introductions to Ireland. For Periods A–F, the number of introductions detected vs. predicted via extrapolation for these periods were 170/186, 41/42, 218/224, 728/867, 1937/2258, and 1309/1383, respectively. Similarly the proportions of sequences available to the number estimated to detect all introductions were 1347/2044, 450/570, 3311/4303, 6536/11815, 11503/19538, and 6371/8751, respectively. Such extrapolations could provide a reliable approach for predicting the total number of introductions and required sequencing levels in comparable epidemiological scenarios (i.e., subject virus or geographic location).

**Table 1.** SARS-CoV-2 introductions detected vs. predicted and sequences identified vs. estimated sequences required for Periods A–F in Ireland

| Period | Introductions detected vs. predicted |  | Sequences identified vs. estimated sequences required |  |
| --- | --- | --- | --- | --- |
|  | Detected | Predicted | Identified | Required |
| A | 170 | 186 | 1347 | 2044 |
| B | 41 | 42 | 450 | 570 |
| C | 218 | 224 | 3311 | 4303 |
| D | 728 | 867 | 6536 | 11815 |
| E | 1937 | 2258 | 11503 | 19538 |
| F | 1309 | 1383 | 6371 | 8751 |

In contrast to Figure 6C&D which refer to the whole island, Figure 6E shows the number of introductions to NI and RoI during Periods A–F. Normalising both introductions and sequences acquired per capita accounts for biases in total population and disparities in sequencing efforts. This reveals that introduction rates per capita were similar between NI (circles) and RoI (squares) with the exception of Period D during which they were higher in NI. This approach also allows comparison of sequencing levels per capita achieved during each period (rightmost points).

### Population density and deprivation significantly impact SARS-CoV-2 introductions and spread in Ireland

We examined the effect of population density and deprivation on the spread of the virus within NI and RoI using six ordinary least-squares (OLS) regression models and Geographic Information Systems (GIS). Considering all regression models (see Table 2), variance inflation factors (VIFs) were below 3,indicating that there is no significant multicollinearity among the independent variables. Furthermore, The Koenker (BP) statistics are all above the 0.05 threshold, which suggests that the models do not suffer from heteroskedasticity, and indicate that applying geographically-weighted regression (GWR) would not yield much additional clarity. Analysis of Periods D–F, which had the highest sequencing rates, revealed that both population density and deprivation had some statistically significant correlations with the introduction and spread of the virus.

**Table 2.** Summary of OLS regression and GIS results for population density and deprivation in Ireland

| Period and country | Variable | Coefficient | Robust $p$ | Adjusted $R^2$ | VIF | Koenker (BP) Statistic |
| --- | --- | --- | --- | --- | --- | --- |
| D NI | Pop. density | $2.4 \times 10^{-5}$ | $6.7 \times 10^{-2}$ | $8.7 \times 10^{-1}$ | 2.8 | 0.38 |
| | Deprivation | $1.5 \times 10^{-2}$ | $*2.4 \times 10^{-3}$ | | 2.8 | 1.7 |
| E NI | Pop. density | $-6.0 \times 10^{-6}$ | $7.3 \times 10^{-1}$ | $5.2 \times 10^{-1}$ | 2.8 | 0.09 |
| | Deprivation | $1.2 \times 10^{-2}$ | $*2.5 \times 10^{-2}$ | | 2.8 | 1.5 |
| F NI | Pop. density | $-2.0 \times 10^{-6}$ | $9.4 \times 10^{-1}$ | $2.7 \times 10^{-1}$ | 2.8 | 0.13 |
| | Deprivation | $8.6 \times 10^{-3}$ | $2.0 \times 10^{-1}$ | | 2.8 | 0.80 |
| D RoI | Pop. density | $2.6 \times 10^{-4}$ | $*\sim 0$ | $9.6 \times 10^{-1}$ | 1.2 | 0.48 |
| | Deprivation | $-2.9 \times 10^{-3}$ | $6.5 \times 10^{-1}$ | | 1.2 | 1.1 |
| E RoI | Pop. density | $8.3 \times 10^{-5}$ | $*\sim 0$ | $3.1 \times 10^{-1}$ | 1.2 | 0.84 |
| | Deprivation | $-2.9 \times 10^{-3}$ | $8.0 \times 10^{-1}$ | | 1.2 | 0.88 |
| F RoI | Pop. density | $1.7 \times 10^{-4}$ | $*\sim 0$ | $8.9 \times 10^{-1}$ | 1.2 | 0.18 |
| | Deprivation | $-1.5 \times 10^{-2}$ | $*2.1 \times 10^{-2}$ | | 1.2 | 1.32 |

Our GIS analysis indicated that the introduction and spread of SARS-CoV-2 infections was not randomly distributed across Ireland. Population density displayed statistically significant positive correlations with SARS-CoV-2 introductions and spread in Period D (p *<* 0.05) in NI and in RoI (p *<* 10^-4^) during all three periods (D–F). More SARS-CoV-2 sequences were produced in more deprived areas of NI in Periods D & E (p *<* 0.05). However, a significant inverse relationship was observed with deprivation during Period F (p *<* 0.05) in RoI. This unexpected result may reflect differing sequencing strategies between NI and RoI. Also, note that these findings are correlational in nature and do not necessarily indicate causation. You may interactively explore our datasets associating population density and deprivation in Ireland to SARS-CoV-2 introductions and spread (http://go.qub.ac.uk/gis-intros-to-ireland).

### The localised SARS-CoV-2 spreading revealed by geospatiotemporal analysis of introduction clusters confirms the validity of our phylogenomic analysis

We conducted geospatiotemporal analyses of introduction clusters by tracking the geographic appearance of phylogenetically-descendent samples linked to each introduction event over time. Samples with incomplete or ambiguous geographic metadata were excluded from geospatial mapping, resulting in 95% (31,529/33,131) of the Irish sequences being retained for analysis. Figure 8A depicts Ireland with local government districts of NI and RoI, while Figure 8B and Figure 8C illustrate the spreading patterns of the third and second-largest detected introduction clusters, respectively. An introduction cluster of 960 Irish Delta SARS-CoV-2 sequences originating from Scotland via Causeway Coast and Glens in NI during Period D (2^nd^ June–31^st^ July 2021) showed intra- and neighbouring-region spread, with a transmission hop from NI to County Roscommon and County Dublin (Figure 8Bii to Figure 8Biii). An even larger introduction cluster of 1,274 Irish Omicron BA.1 SARS-CoV-2 sequences during Period E (30^th^ November 2021–31^st^ January 2022) spread similarly, originating from England via County Dublin and County Kildare (Figure 8C(i-vi)).

**Figure 8.**
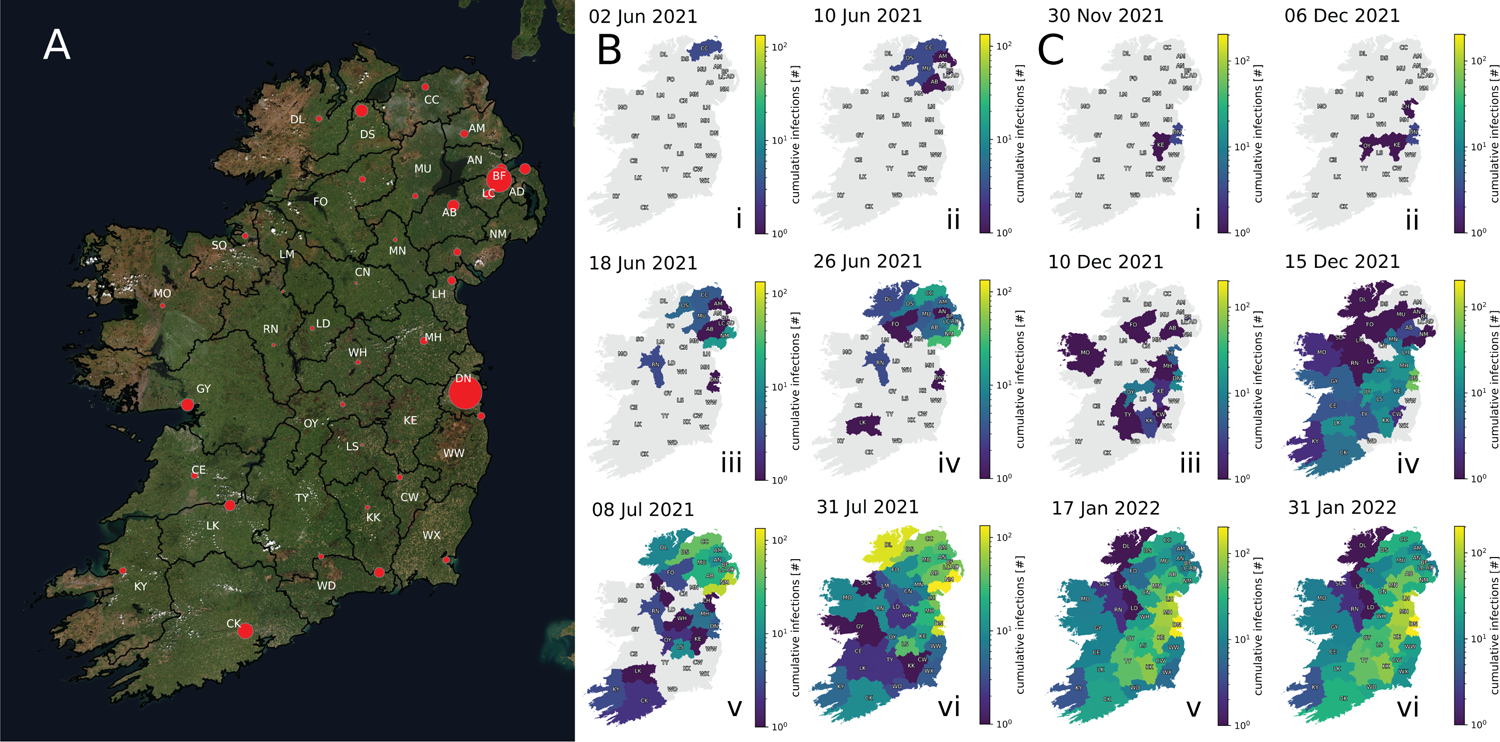
Geospatial tracking of the spread of samples from introduction events within Ireland. **A**, A map of Ireland created with GeoPandas v0.11.1 (Jordahl et al. (2022), https://geopandas.org) using the Esri “World Imagery” basemap (Sources: Esri, DigitalGlobe, GeoEye, i-cubed, USDA FSA, USGS, AEX, Getmapping, Aerogrid, IGN, IGP, swisstopo, and the GIS User Community) as retrieved using contextily v1.2.0 (https://github.com/geopandas/contextily). Red points designate the locations of population centres in each government district with the size of each point scaled according to recent population estimates. Refer to Table S1 for the corresponding metadata for region abbreviations and population centres. **B-i**, An introduction of Delta SARS-CoV-2 descended from Scotland begins an introduction cluster in NI (Causeway Coast and Glens) on 2^nd^ June 2021. **B-ii**, The introduction cluster continues spreading within NI to adjacent and nearby districts, namely Derry and Strabane, Mid and East Antrim, Mid Ulster, Belfast, and Armagh City, Banbridge and Craigavon by 10^th^ June 2021. **B-iii**, The introduction cluster exhibits non-neighbouring spreading events into County Roscommon and County Dublin in RoI as well as adjacently to Lisburn and Castlereagh and Newry, Mourne and Down in NI by 18^th^ June 2021 **B-iv**, The introduction cluster adjacently reaches the remainder of NI and County Donegal in RoI, afflicting Fermanagh and Omagh, Antrim and Newtownabbey, and Ards and North Down as well as hopping to County Limerick in RoI by 26^th^ June 2021. **B-v**, Spreading within NI as well as adjacent spread to additional regions in RoI, namely County Leitrim, County Louth, County Meath, County Westmeath, County Laois, County Kerry, and County Cork occurs by 8^th^ July 2021. **B-vi**, Spreading across Ireland continues throughout July, having touched all government districts in Ireland, afflicting at least 960 individuals within this single viral importation by 31^th^ July 2021. **C-i**, An introduction cluster of BA.1 SARS-CoV-2 descended from England is simultaneously detected within the boundaries of County Dublin and County Kildare in RoI on 30^th^ November 2022. **C-ii**, The introduction cluster adjacently spreads to County Offaly and nearby to County Louth by 6^th^ January 2022. **C-iii**, By 10^th^ December 2021, the cluster rapidly spreads to adjacent districts in RoI, namely County Meath, County Tipperary, County Carlow, and County Kilkenny, and exhibits hops to two districts in NI (Armagh City, Banbridge and Craigavon and Fermanagh and Omagh) and County Mayo in RoI. **C-iv**, The introduction cluster continues rapidly spreading further, reaching Mid Ulster, Antrim and Newtownabbey, Belfast, Armagh City, Banbridge and Craigavon, and Newry, Mourne and Down in NI and County Donegal, County Sligo, County Leitrim, County Longford, County Monaghan, County Westmeath, County Roscommon, County Galway, County Laois, County Clare, County Limerick, County Kerry, and County Cork in RoI by 15^th^ December 2021. **C-v**, SARS-CoV-2 infections linked to this introduction event have reached all districts in Ireland by 17^th^ January 2022. **C-vi**, Spreading across Ireland continues until the last sequenced case in the cluster was recorded on 31^st^ January 2022, reaching at least 1274 individuals within this introduction cluster as tracked over 62 days.

These geospatiotemporal patterns of viral spread are derived solely from phylogenetic trees and associated sample metadata. The fact that they conform to the expected scenario of predominantly local transmission events supports our analytic approach.

Additional regional metadata, including 2-letter abbreviations for local government districts in Figure 8 and information regarding population centres in each district, is provided in Table S1. Supporting Information includes exemplar clusters involving fewer sequences (Figure S10) and animations depicting the introduction and spread of these and the three largest clusters detected (Figures S13–S17) (https://qub-simpson-lab.github.io/SARS-CoV-2-Ireland), including the largest detected cluster of BA.2 SARS-CoV-2 within Period F, which affected 2,377 individuals and lasted 83 days in Ireland (Figure S17).

### Nucleotide substitution rates in Ireland are consistent with global trends

We have assessed SARS-CoV-2 substitution rate over time between the introduced lineages in comparison to reports of others, given the unique geographic and cultural position of Ireland. Our analysis demonstrated that the substitution rates of Irish SARS-CoV-2 were consistent with those earlier reported (Markov et al. 2023; González-Vázquez and Arenas 2023) and were similar to the substitution rates estimated by Nextstrain (Hadfield et al. (2018), https://nextstrain.org).

Figure 9A shows all (as of 18^th^ Feb 2023) Irish sequences with ≥99.9% genome coverage as obtained from GISAID corresponding to lineages introduced in Periods A–F, with the mean observed substitutions tracked for NI and RoI as solid lines. We observe increases at transition periods due to the appearance of new lineages alongside the existing circulating variant (see Markov et al. (2023) for a detailed explanation of this phenomenon). To investigate substitution characteristics of each lineage we separately analysed the apparent substitution rates corresponding to lineages introduced in each period (see Figure S11). Estimated substitution rates for NI and RoI for each lineage group are presented in Figure 9B. From this analysis we can see that overall there do not appear to be significant differences between the apparent substitution rates of SARS-CoV-2 in NI compared to RoI. However, Period B, which is a time period with the lowest sequencing rates both locally and globally (see Figure S12), and Period F, which saw significantly more importations to NI than RoI may be significant.

**Figure 9.**
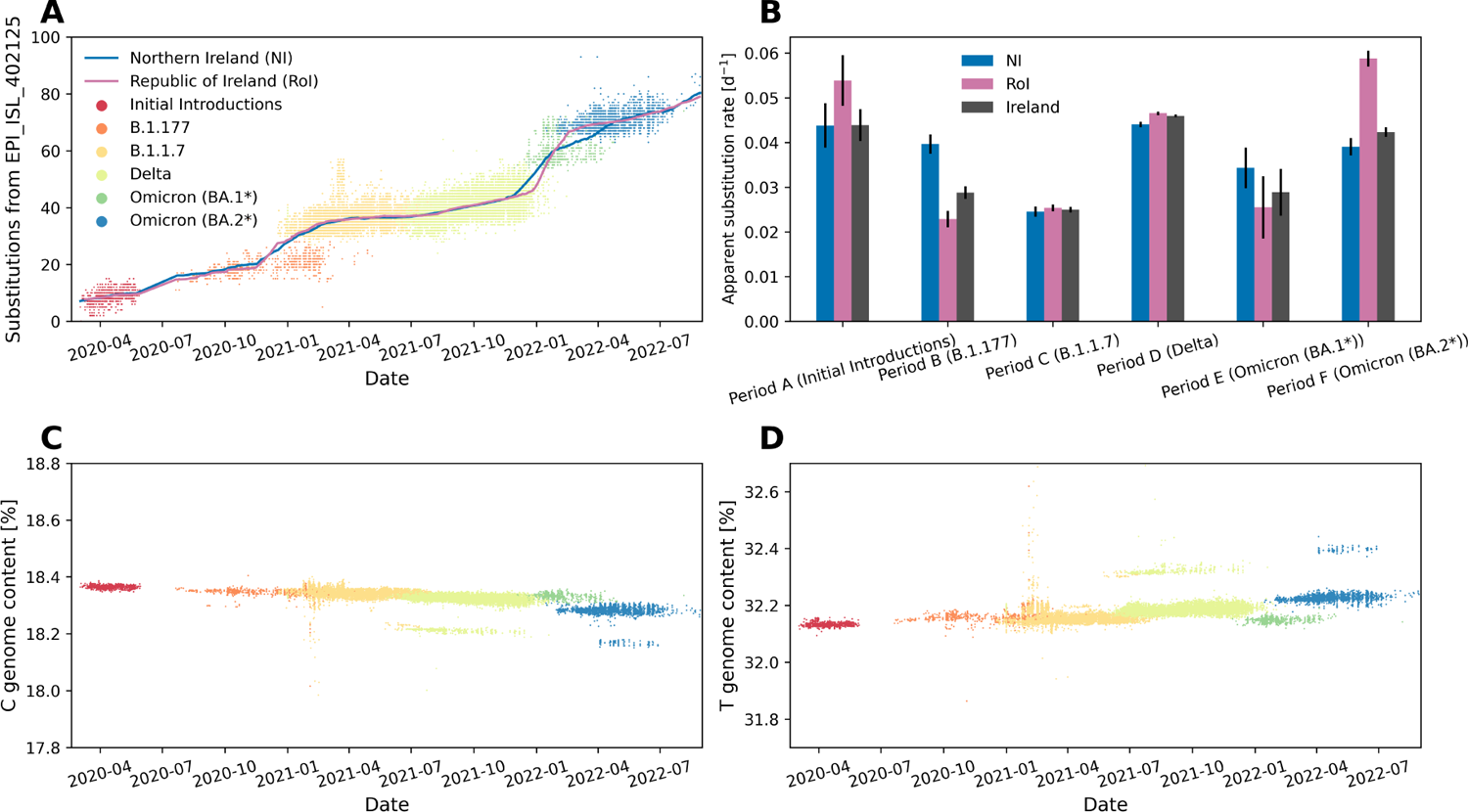
Population-wide SARS-CoV-2 genomic trends in Ireland. Genome coverage for all samples included were at least 99.9% upon alignment to the Wuhan-Hu-1 SARS-CoV-2 reference (GISAID: EPI ISL 402125) **A**, Observed substitutions from the Wuhan-Hu-1 SARS-CoV-2 reference per sequence **B**, Apparent substitution rates per major SARS-CoV-2 lineage related to each studied introduction period. See Figure S11 for a visualisation of each linear regression, and Table S8 for the corresponding statistics. **C**, Genome C content per sequence **D**, Genome T content per sequence.

Furthermore, Figure 9C&D show that the decrease in genome cytosine (C) content (Figure 9C) is generally compensated by an increase in genome thymine (T) content (Figure 9D), which is characteristic of SARS-CoV-2 evolutionary behaviour reported previously (Di Giorgio et al. 2020; Simmonds 2020).

## Discussion

The unprecedented global effort to rapidly sequence millions of SARS-CoV-2 samples and share the results openly has created a rich resource that provides the opportunity to address previously intractable research questions. While the scale of the dataset is its strength, it has posed problems to existing software, tools and methods that had not previously encountered such computational challenges. However, the work of many other research groups to optimise multiple sequence alignment and phylogenetic analysis of millions of SARS-CoV-2 sequences has enabled us to identify introduction events to the island of Ireland during key periods of the pandemic.

The 4,403 independent SARS-CoV-2 introductions to Ireland detected during the six time periods studied should be treated as a conservative lower bound estimate for several reasons. Firstly, the genome sequencing data available is limited by sparseness and a variable sampling rate. In 2020, the rate of sequencing in both NI and RoI was low — with less than three thousand sequences from each, compared to approximately fifty thousand each in 2021. This restricts how informative earlier time periods, specifically A, B and much of C, are for gleaning insights into Irish introductions. Key periods of importation that involve increased international travel and gatherings, such as Christmas, are also periods of reduced sampling. This has the double effect of increasing the rate of successful introductions taking place while also reducing the opportunity for these events to be captured. These factors within the Irish sequencing programmes directly affect the number of introductions observed and cause the extent of underestimation to vary.

Secondly, a global inequality in SARS-CoV-2 genomic surveillance has been reported (Brito et al. 2022). The sampling rate in other countries can affect the number of introductions identified in Ireland. Consider a scenario in which a lineage with a unique set of mutations, lineage L, is circulating in another country, country A. An introduction event from country A to Ireland takes place and if sequenced in Ireland, the unique set of mutations allows us to easily attribute country A as the origin. If a subsequent mutation, mutation M, occurs in country A and this mutated lineage L is again introduced, it will be identified as a second introduction event if sequenced in both countries. If, however, there is a low sequencing rate in country A, mutation M might only be sequenced among Irish samples. Therefore, mutation M would appear as an Ireland-specific mutation occurring within the original introduction cluster of lineage L. While rare, this scenario would result in missed introductions and merged clusters that should be independent from each other. The high rate of sequencing in nearby Great Britain should reduce the occurrence of such scenarios affecting our introduction counts for Ireland.

A low sampling rate in other countries can affect more than just the number of introductions; it can result in introductions being wrongly attributed. For example, scenarios can arise whereby a cryptic lineage circulates in a country, country B, with a low rate of sequencing and is therefore not sampled and attributed as originating in country B. Introductions from country B to other countries than Ireland can also occur, some of which may have robust variant detection programmes, potentially leading to some introductions to Ireland from country B being wrongly attributed to one of these other countries.

Thirdly, to identify independent introductions it is necessary to distinguish them from each other. Genetic diversity enables the tracking of viral infection and onward transmission events for the purposes of genomic surveillance (Grubaugh, Ladner, Lemey, et al. 2018; Rockett et al. 2020). Novel shared mutations reveal a history of infection and transmission events. Similarly, we can only differentiate between separate introduction events if they are genetically distinct. Sequences that are identical but result from multiple introduction events will be grouped in the same introduction cluster because they are indistinguishable. They are therefore counted as a single introduction. The relatively low mutation rate of SARS-CoV-2 compared to other viruses, and subsequently the low level of genetic diversity, will increase the likelihood of this occurring, resulting in an underestimate of independent introduction events.

Notwithstanding the caveats discussed above, we were able to identify patterns of introductions that are not explained by the influence of different national sequencing levels. As expected, for some time periods there is a negative correlation between number of introductions and distance, presumably reflecting less travel from distant countries.

The ability to link many sequences with geospatial and temporal metadata attributable to specific introduction and spreading events enabled us to conduct a range of geospatial analyses. Such analyses provide an opportunity to use geospatiotemporal data to develop epidemiological models and better understand pandemic dynamics. We further hypothesise that, given a sufficiently high level of accurate viral genome sequencing, spatiotemporal clusters of virus spread can be used to predict the presence of undetected introduction events.

The correlations observed between population density, deprivation, and introductions and spread of SARS-CoV-2 suggest roles for these factors in enhancing viral introductions and spread and corroborate previous associations of SARS-CoV-2 incidence with deprivation in Ireland (Madden et al. 2021), particularly in NI (McKinley et al. 2021). Targeted interventions in areas of high population density and deprivation may therefore be warranted to reduce the introductions and spread of SARS-CoV-2 and other future transmissable diseases. However, the relationship between deprivation and geographic location is complex; not all individuals experiencing deprivation live in areas typically associated with deprivation, and not all individuals living in such areas experience deprivation. Thus, incorporating high-resolution geospatial data such as the 890 Super Output Areas (https://www.nisra.gov.uk/support/output-geography-census-2011/super-output-areas) or 3,780 Data Zones (https://www.nisra.gov.uk/support/geography/data-zones-census-2021) in NI compared to the 11 LGDs used in this study, due to the limited resolution of GISAID metadata, could enhance the accuracy of analyses and provide a more nuanced understanding of the distribution of population density and deprivation.

Because multiple factors can influence the number of introductions observed it can be difficult to attribute the cause for differences between time periods. For example, the relative scarcity of introductions observed in Period B is likely due to very limited travel, but is also exacerbated by the low levels of sequencing in Ireland and globally. This raises the salient question: what level of sequencing is required to provide adequate genomic surveillance? Sequencing every single positive case will provide the most complete representation of variants circulating in a community, however, this would be too costly and largely unnecessary as in many cases the same variants would be sampled repeatedly. On the other hand, if too few cases are sequenced, this risks missing variants of concern. These could be imported from elsewhere or arise *de novo* and both would likely not be detected until well established in the community. The optimum number of samples to sequence will vary depending on total number of cases and international sequencing levels. Nonetheless, we have developed a methodology with which to estimate both the total number of importations that are likely to have occurred as well as the minimal number of sequences required to detect all importations for a given time period (see Figure 5). We find that relatively few sequences are required to accurately estimate the total number of introductions (although more are needed as introduction rates increase). However, higher sequencing levels will always help to elucidate infection dynamics and enhance the detection of emerging variants. These factors should be considered when setting sequencing targets.

The higher per capita introduction rate into NI during Period D (Delta) is likely due to higher travel into NI than RoI at this time (Figure 1E). The similar incidence in both jurisdictions during Period D (Figure 1A) suggests that most cases arise due to internal spread and the benefit of travel restrictions is largely limited to reducing the risk of importing novel variants.

To investigate the relationships between viral characteristics (e.g., genetic identity and load) and clinical outcomes (e.g., symptoms, complications, disease severity, duration of illness, and treatment responses), it is necessary to perform viral sequencing of infected individuals. When multiple individual sequences from a sufficiently large proportion of the population are available, this approach enables tracking of viral dynamics through phylogenetic reconstructions, which can identify outbreaks and chains of transmission events. Phylogeographic techniques used in this study allow: 1) estimation of the origin and source of introductions, 2) mapping of subsequent spreading events and propagation, 3) estimation of substitution rates within introduction clusters, and 4) provision of insights into the effects of mutations on viral characteristics. An alternative approach to study population-level variant dynamics is the monitoring of viruses within wastewater using molecular methods. This provides unbiased aggregate sampling of entire communities and its effectiveness has already been demonstrated in Ireland (Wade et al. 2022; Reynolds et al. 2022). Despite the challenges of genome sequencing from wastewater it has been shown to be an effective complementary approach for genomic surveillance of SARS-CoV-2 (Karthikeyan et al. 2022), although without the information that can be gained from the reconstruction of a molecular phylogeny.

In the context of infectious disease surveillance and outbreak response, phylogenetic analyses have shown to be a valuable tool (Attwood et al. 2022) for identifying emerging strains (Tang et al. 2021), tracking the spread of disease (Moreno et al. 2020; Borges et al. 2022), and strategising targeted interventions (Kwon et al. 2021). Incorporating phylogenetic analyses into real-time advisory workflows (Hadfield et al. 2018; Turakhia, Thornlow, A. S. Hinrichs, et al. 2021) requires a robust system for collecting, storing, and analyzing genetic sequence data, as well as trained personnel who are skilled in bioinformatics and phylogenetic analyses (Smith 2018). By leveraging the power of genetic sequencing and analysis, public health agencies and other stakeholders can better understand the evolution and transmission of pathogens, identify at-risk populations, and develop effective strategies to prevent and control outbreaks. Increased use of phylogenetic analyses in real-time advisory workflows has the potential to significantly improve our ability to respond to infectious disease threats, ultimately leading to better public health outcomes.

## Conclusions

Despite the high level of SARS-CoV-2 WGS, particularly in the UK, sampling was relatively sparse and variable. Nonetheless, the potential of genomic epidemiology is well-recognised (Stockdale, Liu, and Colijn 2022; Hill et al. 2021; McBroome et al. 2022; Killough et al. 2023) and this analysis further demonstrates the additional value of WGS beyond surveillance of emerging variants. Although our conclusions must be tempered by fluctuations in local and international sequencing, the estimates of numbers of introductions and geospatial tracking of specific clusters could be used to inform public health agencies how to limit the spread of health risks to neighbouring countries and to prevent unwarranted travel and trade restrictions. These data could also assist vaccine deployment strategies and implicitly estimate herd immunity thresholds (Stockdale, Liu, and Colijn 2022). The goal of the International Health Regulations is to avoid disruption to global travel and trade. Performing this analysis closer to real time and on an international scale could inform policy about the effectiveness of border measures.

The opportunity provided by the continually decreasing cost of WGS should be used to adopt WGS more widely to monitor and geospatiotemporally investigate future infectious conditions. Sequencing all infected individuals is not presently feasible nor is this necessary to predict the total number of introductions. Our findings demonstrate the extensive variation in sequencing requirements across the SARS-CoV-2 pandemic. We therefore recommend a flexible strategy for surveillance with the capability to surge sequencing above baseline rates as required by the contemporaneous conditions.

## Code availability

The analysis of data was conducted in Jupyter Notebook (https://jupyter.org); the analysis notebooks used in this study can be accessed at https://github.com/QUB-Simpson-lab/paper-sars-cov-2-intros-to-ireland.

## Data Availability

GISAID EPI ISL codes for all sequences analysed are provided (https://github.com/QUB-Simpson-lab/paper-sars-cov-2-intros-to-ireland/analysis/datasets/GISAID/sequences.txt). A companion website (https://qub-simpson-lab.github.io/SARS-CoV-2-Ireland), which includes video files S13-S17 showcasing visualizations of geospatial introduction and spreading events of SARS-CoV-2, accompanies this publication. Our datasets associating population density and deprivation in Ireland to SARS-CoV-2 introductions and spread can are available (http://go.qub.ac.uk/gis-intros-to-ireland).

## Acknowledgments

We would like to express our sincere gratitude to several individuals and organizations for their invaluable contributions to this project. Firstly, we would like to acknowledge the GISAID members for providing the database and open access to WGS data, which enabled us to conduct our analyses. We would also like to extend our thanks to COG-UK for their generous funding and provision of sequences, specifically in the context of sequencing in Northern Ireland. Furthermore, we wish to thank the Irish Coronavirus Sequencing Consortium for their critical contributions of sequencing data in the Republic of Ireland. For their invaluable contributions to sequencing NIRE samples during the COVID19 pandemic: Marc-Aurel Fuchs, Julia Miskelly, Courtney Ward, and Chris Baxter of the Genomics Core Technology Unit of Queen’s University Belfast (QUB), Clara Radulescu, Miao Tang, Arun Mariappan, Aditi Singh, Deborah Lavin, Syed Umbreen, Fiona Rogan, Arthur Fitzgerald, and Michael Glenn of the Wellcome-Wolfson Institute for Experimental Medicine at QUB and James McKenna, Eiĺıs McColgan, and Zoltan Molnar of the Regional Virus Laboratory, Belfast Health and Social Care Trust. We are grateful to Theo Sanderson, Richard Goater, Harald Vöhringer, and Jeffery Barrett for developing the covince open-source visualisation platform (https://github.com/covince/covince). We extend thanks to Conor Graham and Emma Delaney at Queen’s University Belfast School of Built Environment for their support in the development and deployment of our fork of the covince platform as hosted at http://go.qub.ac.uk/covid19.

## Author contributions

AMR, EPT, TS, and DAS conceived the study. AMR, EPT, and SB performed bioinformatic processing of SARS-CoV-2 WGS data. AMR developed the downstream analyses of the global SARS-CoV-2 viral phylogeny to prune the data into periods with which to detect introduction clusters. EPT designed geospatiotemporal analyses of SARS-CoV-2 introductions and spread from the phylogeny. AMR and EPT prepared bespoke computer code for the study and will manage the GitHub repository (https://github.com/QUB-Simpson-lab/paper-sars-cov-2-intros-to-ireland). EPT and TMR curated a GitHub Page with a collection of videos showcasing exemplar geospatiotemporal spreading events of SARS-CoV-2 in Ireland (https://qub-simpson-lab.github.io/SARS-CoV-2-Ireland) for the Supporting Information. BFN and JMK assessed the impacts population density and deprivation have on SARS-CoV-2 introductions and spread in Ireland. BFN assembled the ARCGIS website for interactively exploring the geospatial OLS analysis population density and deprivation (http://go.qub.ac.uk/gis-intros-to-ireland). AMR and EPT drafted the paper. AMR, EPT, and BFN created graphical figures to represent the results. The named members of the National SARS-CoV-2 Surveillance & Whole Genome Sequencing (WGS) Programme were responsible for many of the sequences collected in the Republic of Ireland and provided verification of assumptions regarding aspects in discussion of the RoI sequencing programme. COG-UK provided substantial sequencing data and sequencing support for the entirety of the UK. DTB and DF were consulted to evaluate the study’s potential impact on the public health landscape in Ireland. EPT, AMR, SB, DTB, DF, CGGB, TS, and DAS critically evaluated the manuscript, with all authors approving of its published form.

## References

Alpert, Tara et al. (May 2021). “Early introductions and transmission of SARS-CoV-2 variant B.1.1.7 inthe United States”. In: Cell 184.10, 2595–2604.e13. doi: 10.1016/j.cell.2021.03.061.

Amicone, Massimo, et al. (Mar. 2022). “Mutation rate of SARS-CoV-2 and emergence of mutators during experimental evolution”. In: Evolution, Medicine, and Public Health 10.1, pp. 142–155. doi: 10.1093/emph/eoac010.

Andersen, Kristian G., et al. (Mar. 2020). “The proximal origin of SARS-CoV-2”. In: Nature Medicine 26.4, pp. 450–452. doi: 10.1038/s41591-020-0820-9.

Attwood, Stephen W., et al. (Apr. 2022). “Phylogenetic and phylodynamic approaches to understanding and combating the early SARS-CoV-2 pandemic”. In: Nature Reviews Genetics 23.9, pp. 547–562. doi: 10.1038/s41576-022-00483-8.

Bach-Mortensen, Anders Malthe and Michelle Degli Esposti (Feb. 2021). “Is area deprivation associated with greater impacts of COVID-19 in care homes across England? A preliminary analysis of COVID-19 outbreaks and deaths”. In: Journal of Epidemiology & Community Health 75.7, pp. 624–627. doi: 10.1136/jech-2020-215039. eprint: https://jech.bmj.com/content/75/7/624.full.pdf.

Baillie, Gregory J., et al. (Jan. 2012). “Evolutionary Dynamics of Local Pandemic H1N1/2009 Influenza Virus Lineages Revealed by Whole-Genome Analysis”. In: Journal of Virology 86.1, pp. 11–18. doi: 10.1128/JVI.05347-11.

Bedford, Trevor, et al. (Sept. 2020). “Cryptic transmission of SARS-CoV-2 in Washington state”. In: Science 370.6516, pp. 571–575. doi: 10.1126/science.abc0523. eprint: https://www.science.org/doi/pdf/10.1126/science.abc0523.

Bondarenko, Maksym, et al. (Sept. 2020). Census/projection-disaggregated gridded population datasets for 189 countries in 2020 using Built-Settlement Growth Model (BSGM) outputs. University of Southampton. doi: 10.5258/SOTON/WP00684.

Borges, Vítor, et al. (Jan. 2022). “SARS-CoV-2 introductions and early dynamics of the epidemic in Portugal”. In: Communications Medicine 2, p. 10. doi: 10.1038/s43856-022-00072-0.

Brian, David A. and Ralph S. Baric (Oct. 2005). “Coronavirus Genome Structure and Replication”. In: *Coronavirus Replication and Reverse Genetics*. Ed. by Luis Enjuanes. Berlin, Heidelberg: Springer Berlin Heidelberg, pp. 1–30. isbn: 978-3-540-26765-2. doi: 10.1007/3-540-26765-4_1.

Brito, Anderson F., et al. (Nov. 2022). “Global disparities in SARS-CoV-2 genomic surveillance”. In: Nature Communications 13.1, p. 7003. doi: 10.1038/s41467-022-33713-y.

Bukhari, Ali Raza, et al. (Mar. 2023). “Sequential viral introductions and spread of BA.1 drove the Omicron wave across Pakistani provinces”. In: medRxiv. doi: 10.1101/2023.03.25.23287718. eprint: https://www.medrxiv.org/content/early/2023/03/27/2023.03.25.23287718.full.pdf.

Castillo-Morales, Atahualpa, et al. (Oct. 2021). “Causes and consequences of purifying selection on sars-cov-2”. In: Genome Biology and Evolution 13.10. Ed. by Mario dos Reis, evab196. doi: 10.1093/gbe/evab196.

da Silva Filipe, Ana, et al. (Jan. 2021). “Genomic epidemiology reveals multiple introductions of SARS-CoV-2 from mainland Europe into Scotland”. In: Nature Microbiology 6.1, pp. 112–122. doi: 10.1038/s41564-020-00838-z.

Deng, Xianding et al. (June 2020). “Genomic surveillance reveals multiple introductions of SARS-CoV-2 into Northern California”. In: Science 369.6503, pp. 582–587. doi: 10.1126/science.abb9263. eprint: https://www.science.org/doi/pdf/10.1126/science.abb9263.

Di Giorgio, Salvatore, et al. (June 2020). “Evidence for host-dependent RNA editing in the transcriptome of SARS-CoV-2”. In: Science Advances 6.25, eabb5813. doi: 10.1126/sciadv.abb5813. eprint: https://www.science.org/doi/pdf/10.1126/sciadv.abb5813.

Díez-Fuertes, Francisco, et al. (Apr. 2020). “Phylodynamics of SARS-CoV-2 transmission in Spain”. In: *bioRxiv*. doi: 10.1101/2020.04.20.050039. eprint: https://www.biorxiv.org/content/early/2020/04/20/2020.04.20.050039.full.pdf.

du Plessis, Louis, et al. (Feb. 2021). “Establishment and lineage dynamics of the SARS-CoV-2 epidemic in the UK”. In: Science 371.6530, pp. 708–712. doi: 10.1126/science.abf2946.

Duchene, Sebastian et al. (July 2020). “Temporal signal and the phylodynamic threshold of SARS-CoV-2”. In: Virus Evolution 6.2. doi: 10.1093/ve/veaa061.

Dudas, Gytis, et al. (Apr. 2017). “Virus genomes reveal factors that spread and sustained the Ebola epidemic”. In: Nature 544.7650, pp. 309–315. doi: 10.1038/nature22040.

Elliott, Paul et al. (June 2022). “Twin peaks: The Omicron SARS-CoV-2 BA.1 and BA.2 epidemics inEngland”. In: Science 376.6600, eabq4411. doi: 10.1126/science.abq4411.

Firoozi Nejad, Behnam (June 2016). “Population mapping using census data, GIS and remote sensing”. https://ethos.bl.uk/OrderDetails.do?uin=uk.bl.ethos.705917. PhD thesis. Faculty of Engineering and Physical Sciences, Chapter 5.

Fonager, Jannik, et al. (Mar. 2022). “Molecular epidemiology of the SARS-CoV-2 variant Omicron BA.2 sub-lineage in Denmark, 29 November 2021 to 2 January 2022”. In: Eurosurveillance 27.10. doi: 10.2807/1560-7917.ES.2022.27.10.2200181.

Fuchs, Marc, et al. (Mar. 2022). “Mini-XT, a miniaturized tagmentation-based protocol for efficient sequencing of SARS-CoV-2”. In: Journal of Translational Medicine 20.1, p. 105. doi: 10.1186/s12967-022-03307-9.

Gámbaro, Fabiana, et al. (July 2020). “Introductions and early spread of SARS-CoV-2 in France, 24 January to 23 March 2020”. In: Eurosurveillance 25.26, 2001200. doi: 10.2807/1560-7917.ES.2020.25.26.2001200.

Githinji, George, et al. (Aug. 2021). “Tracking the introduction and spread of SARS-CoV-2 in coastal Kenya”. In: Nature Communications 12.1, p. 4809. doi: 10.1038/s41467-021-25137-x.

Gonzalez-Reiche, Ana S. et al. (May 2020). “Introductions and early spread of SARS-CoV-2 in the New York City area”. In: Science 369.6501, pp. 297–301. doi: 10.1126/science.abc1917. eprint: https://www.science.org/doi/pdf/10.1126/science.abc1917.

Gonźalez-Vázquez, Luis Daniel and Miguel Arenas (Feb. 2023). “Molecular Evolution of SARS-CoV-2 during the COVID-19 Pandemic”. In: Genes 14.2. doi: 10.3390/genes14020407.

Grubaugh, Nathan D., Jason T. Ladner, Moritz U. G. Kraemer, et al. (June 2017). “Genomic epidemiology reveals multiple introductions of Zika virus into the United States”. In: Nature 546.7658, pp. 401–405. doi: 10.1038/nature22400.

Grubaugh, Nathan D., Jason T. Ladner, Philippe Lemey, et al. (Dec. 2018). “Tracking virus outbreaks in the twenty-first century”. In: Nature Microbiology 4.1, pp. 10–19. doi: 10.1038/s41564-018-0296-2.

Hadfield, James et al. (May 2018). “Nextstrain: real-time tracking of pathogen evolution”. In: Bioinformatics 34.23, pp. 4121–4123. doi: 10.1093/bioinformatics/bty407. eprint: https://academic.oup.com/bioinformatics/article-pdf/34/23/4121/48921142/bioinformatics\_34\_23\_4121.pdf.

Hamming, R. W. (Apr. 1950). “Error detecting and error correcting codes”. In: The Bell System Technical Journal 29.2, pp. 147–160. doi: 10.1002/j.1538-7305.1950.tb00463.x.

Harvey, William T. et al. (June 2021). “SARS-CoV-2 variants, spike mutations and immune escape”. In: Nature Reviews Microbiology 19.7. (COG-UK), pp. 409–424. doi: 10.1038/s41579-021-00573-0.

Hill, Verity, et al. (Dec. 2021). “Progress and challenges in virus genomic epidemiology”. In: Trends in Parasitology 37.12. Publisher: Elsevier, pp. 1038–1049. doi: 10.1016/j.pt.2021.08.007.

Ishikawa, Sohta A. et al. (May 2019). “A Fast Likelihood Method to Reconstruct and Visualize Ancestral Scenarios”. In: Molecular Biology and Evolution 36.9, pp. 2069–2085. doi: 10.1093/molbev/msz131.

Jordahl, Kelsey et al. (July 2022). *geopandas/geopandas: v0.11.1*. Version v0.11.1. doi: 10.5281/zenodo.6894736.

Juscamayta-Ĺopez, Eduardo, et al. (July 2021). “Phylogenomics reveals multiple introductions and early spread of SARS-CoV-2 into Peru”. In: Journal of Medical Virology 93.10, pp. 5961–5968. doi: 10.1002/jmv.27167. eprint: https://onlinelibrary.wiley.com/doi/pdf/10.1002/jmv.27167.

Karthikeyan, Smruthi et al. (July 2022). “Wastewater sequencing reveals early cryptic SARS-CoV-2 variant transmission”. In: Nature 609.7925, pp. 101–108. doi: 10.1038/s41586-022-05049-6.

Katoh, Kazutaka et al. (July 2002). “MAFFT: a novel method for rapid multiple sequence alignment based on fast Fourier transform”. In: Nucleic Acids Research 30.14, pp. 3059–3066. doi: 10.1093/nar/gkf436. eprint: https://academic.oup.com/nar/article-pdf/30/14/3059/9488148/gkf436.pdf.

Kenny, Kevin (June 2014). The American Irish: A History. Routledge. isbn: 9781315842530. doi: 10.4324/9781315842530.

Khare, Shruti, et al. (Dec. 2021). “GISAID’s Role in Pandemic Response”. In: China CDC Weekly 3.49, pp. 1049–1051. doi: 10.46234/ccdcw2021.255.

Killough, Nicholas, et al. (Feb. 2023). “How public health authorities can use pathogen genomics inhealth protection practice: a consensus-building Delphi study conducted in the United Kingdom”. In: Microbial Genomics 9.2, 000912. doi: 10.1099/mgen.0.000912.

Komissarov, Andrey B., et al. (Jan. 2021). “Genomic epidemiology of the early stages of the SARS-CoV-2 outbreak in Russia”. In: Nature Communications 12.1, p. 649. doi: 10.1038/s41467-020-20880-z.

Kwon, Jung-Hoon, et al. (Sept. 2021). “Genomic epidemiology reveals the reduction of the introduction and spread of SARS-CoV-2 after implementing control strategies in Republic of Korea, 2020”. In: Virus Evolution 7.2. veab077. doi: 10.1093/ve/veab077. eprint: https://academic.oup.com/ve/article-pdf/7/2/veab077/50057733/veab077.pdf.

Lanfear, Rob (Oct. 2020). A global phylogeny of SARS-CoV-2 sequences from GISAID. doi: 10.5281/zenodo.3958883.

Lemieux, Jacob E., et al. (Dec. 2021). “Phylogenetic analysis of SARS-CoV-2 in Boston highlights the impact of superspreading events”. In: Science 371.6529, eabe3261. doi: 10.1126/science.abe3261. eprint: https://www.science.org/doi/pdf/10.1126/science.abe3261.

Lemoine, Fŕedéric and Olivier Gascuel (Aug. 2021). “Gotree/Goalign: toolkit and Go API to facilitate the development of phylogenetic workflows”. In: NAR Genomics and Bioinformatics 3.3. doi: 10.1093/nargab/lqab075.

Li, Xiaojun et al. (July 2020). “Emergence of SARS-CoV-2 through recombination and strong purifying selection”. In: Science Advances 6.27, eabb9153. doi: 10.1126/sciadv.abb9153. eprint: https://www.science.org/doi/pdf/10.1126/sciadv.abb9153.

Lobiuc, Andrei et al. (July 2021). “Introduction and Characteristics of SARS-CoV-2 in North-East of Romania During the First COVID-19 Outbreak”. In: Frontiers in Microbiology 12, p. 654417. doi: 10.3389/fmicb.2021.654417.

Lone, Nazir I., et al. (Feb. 2021). “Influence of socioeconomic deprivation on interventions and outcomes for patients admitted with COVID-19 to critical care units in Scotland: A national cohort study”. In: The Lancet Regional Health - Europe 1, p. 100005. doi: https://doi.org/10.1016/j.lanepe.2020.100005.

Madden, Jamie M. et al. (June 2021). “Population Mobility Trends, Deprivation Index and the Spatio-Temporal Spread of Coronavirus Disease 2019 in Ireland”. In: International Journal of Environmental Research and Public Health 18.12. doi: 10.3390/ijerph18126285.

Mallon, Patrick W. G., et al. (Feb. 2021). “Whole-genome sequencing of SARS-CoV-2 in the Republic of Ireland during waves 1 and 2 of the pandemic”. In: medRxiv, p. 2021.02.09.21251402. doi: 10.1101/2021.02.09.21251402.

Markov, Peter V., et al. (Apr. 2023). “The evolution of SARS-CoV-2”. In: Nature Reviews Microbiology. Publisher: Nature Publishing Group, pp. 1–19. doi: 10.1038/s41579-023-00878-2.

Maurano, Matthew T., et al. (Dec. 2020). “Sequencing identifies multiple early introductions of SARS-CoV-2 to the New York City region”. In: Genome Research 30.12, pp. 1781–1788. doi: 10.1101/gr.266676.120.

McBroome, Jakob et al. (June 2022). “Identifying SARS-CoV-2 regional introductions and transmission clusters in real time”. In: Virus Evolution 8.1. veac048. doi: 10.1093/ve/veac048. eprint: https://academic.oup.com/ve/article-pdf/8/1/veac048/49236267/veac048.pdf.

McKinley, Jennifer M et al. (June 2021). “Association between community-based self-reported COVID-19 symptoms and social deprivation explored using symptom tracker apps: a repeated cross-sectional study in Northern Ireland”. In: BMJ Open 11.6. doi: 10.1136/bmjopen-2020-048333. eprint: https://bmjopen.bmj.com/content/11/6/e048333.full.pdf.

Michaelsen, Thomas Y. et al. (May 2022). “Introduction and transmission of SARS-CoV-2 lineage B.1.1.7, Alpha variant, in Denmark”. In: Genome Med. 14.1, p. 47. doi: 10.1186/s13073-022-01045-7.

Moreno, Gage K., et al. (Nov. 2020). “Revealing fine-scale spatiotemporal differences in SARS-CoV-2 introduction and spread”. In: Nature Communications 11.1, p. 5558. doi: 10.1038/s41467-020-19346-z.

Muenchhoff, Maximilian, et al. (Oct. 2021). “Genomic epidemiology reveals multiple introductions of SARS-CoV-2 followed by community and nosocomial spread, Germany, February to May 2020”. In: Eurosurveillance 26.43, 2002066. doi: 10.2807/1560-7917.ES.2021.26.43.2002066.

Murall, Carmen Ĺıa, et al. (Oct. 2021). “A small number of early introductions seeded widespread transmission of SARS-CoV-2 in Qúebec, Canada”. In: Genome Medicine 13.1, p. 169. doi: 10.1186/s13073-021-00986-9.

Nasir, Asghar, et al. (Mar. 2022). “Evolutionary history and introduction of SARS-CoV-2 Alpha VOC/B.1.1.7 in Pakistan through international travelers”. In: Virus Evolution 8.1, veac020. doi: 10.1093/ve/veac020.

Northern Ireland Statistics and Research Agency (Jan. 2013). SOA Background Paper. — (Nov. 2017). Northern Ireland Multiple Deprivation Measure 2017 (NIMDM2017) - SOA level results. O’Connor, Thomas and Mary Ann Lyons (Jan. 2003). Irish migrants in Europe after Kinsale, 1602–1820. Four Courts Press. isbn: 1-85182-701-3.

O’Toole, Á ine, et al. (Feb. 2022). “Pango lineage designation and assignment using SARS-CoV-2 spike gene nucleotide sequences”. In: BMC Genomics 23.1, p. 121. doi: 10.1186/s12864-022-08358-2.

O’Toole, Á ine, et al. (July 2021). “Assignment of epidemiological lineages in an emerging pandemic using the pangolin tool”. In: Virus Evolution 7.2. doi: 10.1093/ve/veab064.

Office for National Statistics (June 2020). Deaths involving COVID-19 by local area and socioeconomic deprivation: deaths occurring between 1 March and 31 July 2020. UK Statistics Authority.

Pachetti, Maria, et al. (Apr. 2020). “Emerging SARS-CoV-2 mutation hot spots include a novel RNA-dependent-RNA polymerase variant”. In: Journal of Translational Medicine 18.1, p. 179. doi: 10.1186/s12967-020-02344-6.

Paiva, Marcelo Henrique Santos, et al. (Dec. 2020). “Multiple Introductions Followed by Ongoing Community Spread of SARS-CoV-2 at One of the Largest Metropolitan Areas of Northeast Brazil”. In: Viruses 12.12. doi: 10.3390/v12121414.

Pedregosa, Fabian, et al. (Nov. 2011). “Scikit-Learn: Machine Learning in Python”. In: *J*. *Mach. Learn. Res*. 12.null, pp. 2825–2830.

Public Health England (June 2020). COVID-19: review of disparities in risks and outcomes. GOV.UK, PHE publications gateway number: GW–1447.

Reynolds, Liam J., et al. (Sept. 2022). “SARS-CoV-2 variant trends in Ireland: Wastewater-based epidemiology and clinical surveillance”. In: Science of The Total Environment 838, p. 155828. doi: 10.1016/j.scitotenv.2022.155828.

Rockett, Rebecca J., et al. (Sept. 2020). “Revealing COVID-19 transmission in Australia by SARS-CoV-2 genome sequencing and agent-based modeling”. In: Nature Medicine 26.9, pp. 1398–1404. doi: 10.1038/s41591-020-1000-7.

Sanderson, Theo (July 2022). “Chronumental: time tree estimation from very large phylogenies”. In: *bioRxiv*, p. 2021.10.27.465994. doi: 10.1101/2021.10.27.465994.

Seabold, Skipper and Josef Perktold (2010). “statsmodels: Econometric and statistical modeling with python”. In: 9th Python in Science Conference.

Simmonds, P. (June 2020). “Rampant C→U Hypermutation in the Genomes of SARS-CoV-2 and Other Coronaviruses: Causes and Consequences for Their Short- and Long-Term Evolutionary Trajectories”. In: mSphere 5.3, e00408–20. doi: 10.1128/mSphere.00408-20. eprint: https://journals.asm.org/doi/pdf/10.1128/mSphere.00408-20.

Smith, David R (May 2018). “Bringing bioinformatics to the scientific masses”. In: EMBO reports 19.6, e46262. doi: 10.15252/embr.201846262. eprint: https://www.embopress.org/doi/pdf/10.15252/embr.201846262.

Stefanelli, Paola, et al. (Apr. 2020). “Whole genome and phylogenetic analysis of two SARS-CoV-2 strains isolated in Italy in January and February 2020: additional clues on multiple introductions and further circulation in Europe”. In: Eurosurveillance 25.13, 2000305. doi: 10.2807/1560-7917.ES.2020.25.13.2000305.

Stockdale, Jessica E., Pengyu Liu, and Caroline Colijn (Nov. 2022). “The potential of genomics for infectious disease forecasting”. In: Nature Microbiology 7.11. Number: 11 Publisher: Nature Publishing Group, pp. 1736–1743. doi: 10.1038/s41564-022-01233-6.

Sukumaran, Jeet and Mark T. Holder (June 2010). “DendroPy: a Python library for phylogenetic computing”. In: Bioinformatics 26.12, pp. 1569–1571. doi: 10.1093/bioinformatics/btq228.

Swofford, David L. and Wayne P. Maddison (Dec. 1987). “Reconstructing ancestral character states under Wagner parsimony”. In: Mathematical Biosciences 87.2, pp. 199–229. doi: 10.1016/0025-5564(87)90074-5.

Talic, Stella, et al. (Oct. 2021). “Effectiveness of public health measures in reducing the incidence ofcovid-19, SARS-CoV-2 transmission, and covid-19 mortality: systematic review and meta-analysis”. In: BMJ 375, e068302. doi: 10.1136/bmj-2021-068302.

Tang, Julian W., et al. (Jan. 2021). “Introduction of the South African SARS-CoV-2 variant 501Y.V2 into the UK”. In: Journal of Infection 82.4. doi: 10.1016/j.jinf.2021.01.007, e8–e10. doi: 10.1016/j.jinf.2021.01.007.

Teljeur, Conor, et al. (Nov. 2019). The Trinity National Deprivation Index for Health and Health Services Research 2016.

The COVID-19 Genomics UK (COG-UK) Consortium (June 2020). “An integrated national scale SARS-CoV-2 genomic surveillance network”. In: The Lancet Microbe 1.3. doi: 10.1016/S2666-5247(20)30054-9, e99–e100. doi: 10.1016/S2666-5247(20)30054-9.

Toovey, Oliver T. R., et al. (Feb. 2021). “Introduction of Brazilian SARS-CoV-2 484K.V2 related variants into the UK”. In: Journal of Infection 82.5. doi: 10.1016/j.jinf.2021.01.025, e23–e24. doi: 10.1016/j.jinf.2021.01.025.

Tordoff, Diana M., et al. (Sept. 2021). “Phylogenetic estimates of SARS-CoV-2 introductions into Washington State”. In: The Lancet Regional Health - Americas 1, p. 100018. doi: 10.1016/j.lana.2021.100018.

Turakhia, Yatish, Bryan Thornlow, Angie Hinrichs, et al. (Aug. 2022). “Pandemic-scale phylogenomics reveals the SARS-CoV-2 recombination landscape”. In: Nature 609.7929, pp. 994–997. doi: 10.1038/s41586-022-05189-9.

Turakhia, Yatish, Bryan Thornlow, Angie S. Hinrichs, et al. (May 2021). “Ultrafast Sample placement on Existing tRees (UShER) enables real-time phylogenetics for the SARS-CoV-2 pandemic”. In: Nature Genetics 53.6, pp. 809–816. doi: 10.1038/s41588-021-00862-7.

Virological (Jan. 2020). Novel 2019 coronavirus genome. url: https://virological.org/t/novel-2019-coronavirus-genome/319.

Virtanen, Pauli, et al. (Mar. 2020). “SciPy 1.0: Fundamental Algorithms for Scientific Computing in Python”. In: Nature Methods 17, pp. 261–272. doi: 10.1038/s41592-019-0686-2.

Wade, Matthew J., et al. (Feb. 2022). “Understanding and managing uncertainty and variability for wastewater monitoring beyond the pandemic: Lessons learned from the United Kingdom national COVID-19 surveillance programmes”. In: Journal of Hazardous Materials 424, p. 127456. doi: 10.1016/j.jhazmat.2021.127456.

Wang, Qian et al. (May 2022). “Antibody evasion by SARS-CoV-2 Omicron subvariants BA.2.12.1, BA.4 and BA.5”. In: Nature 608.7923, pp. 603–608. doi: 10.1038/s41586-022-05053-w.

