## Supporting Information for "SARS-CoV-2 introductions to the island of Ireland"

[1] Wellcome–Wolfson Institute for Experimental Medicine, School of Medicine, Dentistry and Biomedical Sciences, Queen's University Belfast, Belfast, Northern Ireland, BT9 7BL, United Kingdom
[2] Geography, School of Natural and Built Environment, Queen's University Belfast, Belfast, Northern Ireland, BT7 1NN, United Kingdom
[3] The full list of all individual members and partners of COG-UK can be found at `https://www.cogconsortium.uk/full-list-of-all-individual-members-and-partners-of-cog-uk`
[4] Named programme members: Michael J. Carr, Gabriel Gonzalez, Jonathan Dean, Daniel Hare, and Cillian F. De Gascun. National Virus Reference Laboratory (NVRL), School of Medicine, University College Dublin 4, Belfield, D04 V1W8, Dublin, Ireland
[5] Public Health Agency, Health and Social Care Northern Ireland, Belfast, Northern Ireland, BT2 8BS, United Kingdom
[6] Centre for Public Health, School of Medicine, Dentistry and Biomedical Sciences, Queen's University Belfast, Belfast, Northern Ireland, BT12 6BA, United Kingdom
[7] Regional Virus Laboratory, Belfast Health and Social Care Trust, Belfast, Northern Ireland, BT12 6BA, United Kingdom
[8] Institute for Global Food Security, School of Biological Sciences, Queen's University Belfast, Belfast, Northern Ireland, BT9 5DL, United Kingdom
[9] Medical Biology Centre, School of Pharmacy, Queen's University Belfast, Belfast, Northern Ireland, BT9 7BL, United Kingdom

[†] The authors consider these individuals to be Joint First Authors.
[‡] Current address: Milner Centre for Evolution, Department of Life Sciences, University of Bath, Claverton Down, Bath, England, BA2 7AZ, United Kingdom

### List of figures

Figure S1 – GISAID global SARS-CoV-2 phylogeny
Figure S2 – Sample collection date from metadata and their predicted date from time-tree estimate
Figure S3 – Schematic phylogeny
Figure S4 – Period A pruned tree
Figure S5 – Period B pruned tree
Figure S6 – Period C pruned tree
Figure S7 – Period D pruned tree
Figure S8 – Period E pruned tree
Figure S9 – Period F pruned tree
Figure S10 – Additional examples of geospatial spreading of introduced SARS-CoV-2 infection clusters
Figure S11 – OLS regressions of substitutions in Irish SARS-CoV-2 sequences over time per major introduction lineage
Figure S12 – World maps depicting the ratio of introductions to Ireland given the proportion of samples in the global phylogeny for each period.
Figures S13–S17 – Animations depicting geospatiotemporal spread for exemplar SARS-CoV-2 introduction events can be found here.

### List of tables

Table S1 – Metadata for mapping samples to local government districts in Ireland (NI and RoI)
Table S2 – Originating countries and their frequencies of importations for Period A
Table S3 – Originating countries and their frequencies of importations for Period B
Table S4 – Originating countries and their frequencies of importations for Period C

Table S5 – Originating countries and their frequencies of importations for Period D
Table S6 – Originating countries and their frequencies of importations for Period E
Table S7 – Originating countries and their frequencies of importations for Period F
Table S8 – OLS linear regression statistics for estimating substitution rates of major imported lineages to Ireland

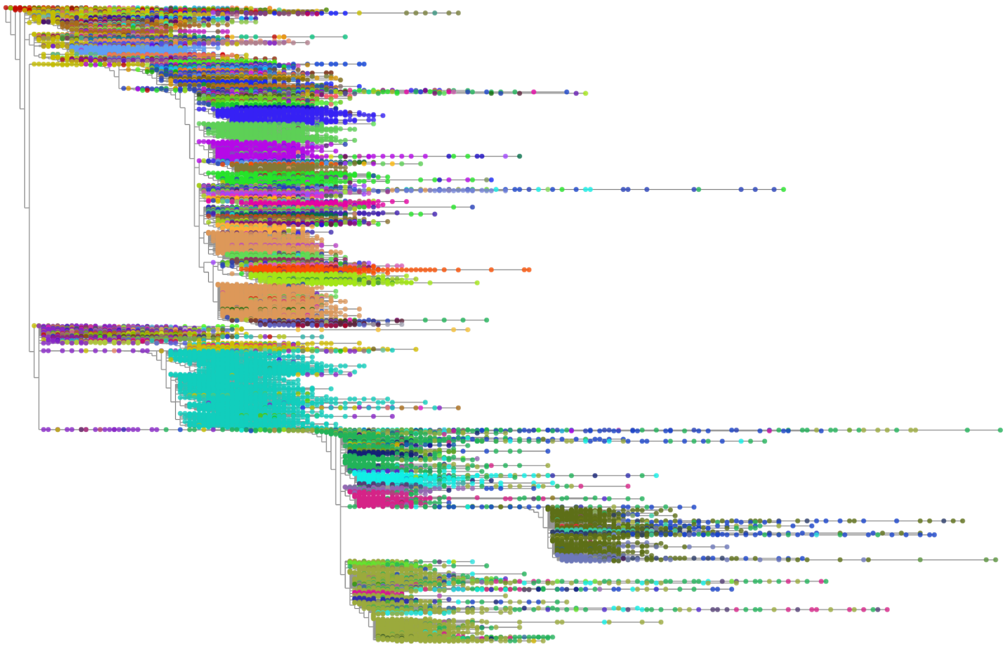

**Figure S1. GISAID global SARS-CoV-2 phylogeny** Global SARS-CoV-2 phylogeny, dated 20$^{th}$ May 2022, from GISAID of 7,603,547 SARS-CoV-2 genome sequences. Samples coloured by Pango (O'Toole et al. 2022) lineage and visualised using Taxonium (Sanderson 2022).

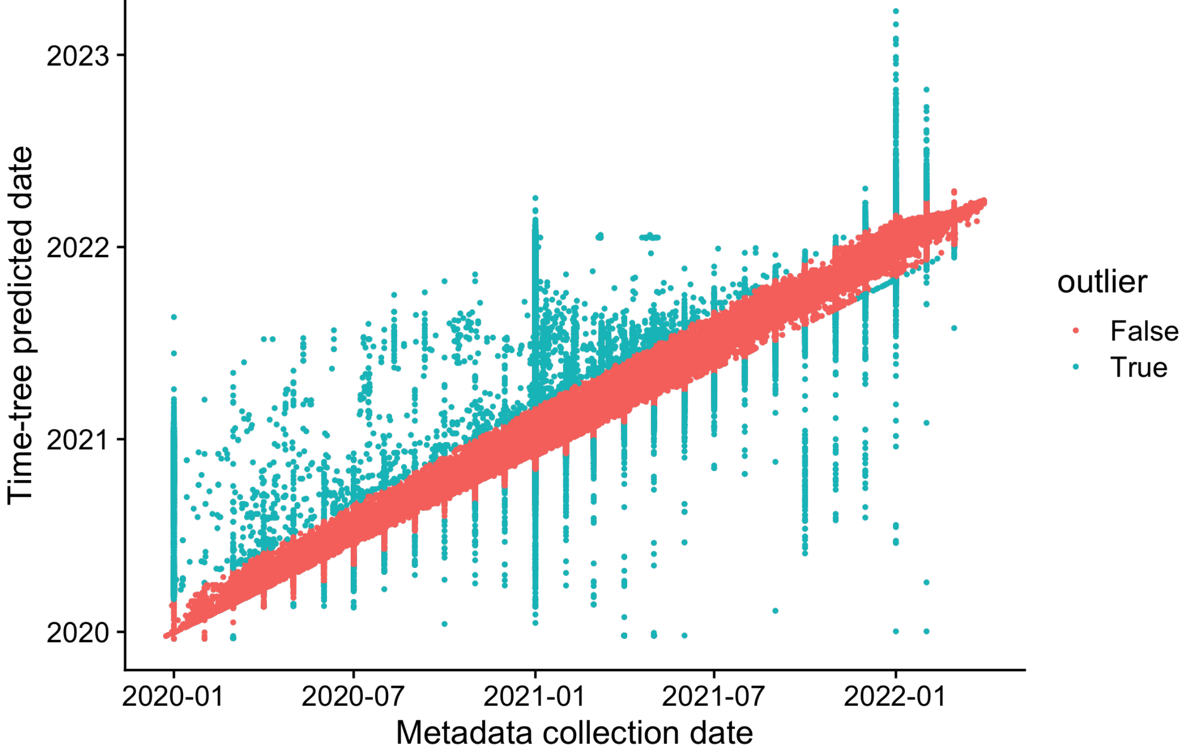


**Figure S2. Sample collection date from metadata and their predicted date from time-tree estimate** Sample collection date from GISAID metadata of sequences in phylogenetic tree and their predicted date from Chronumental time-tree estimate. Sequences that have a difference between metadata date and predicted date of Z-score greater ± 3 are highlighted here as outliers.

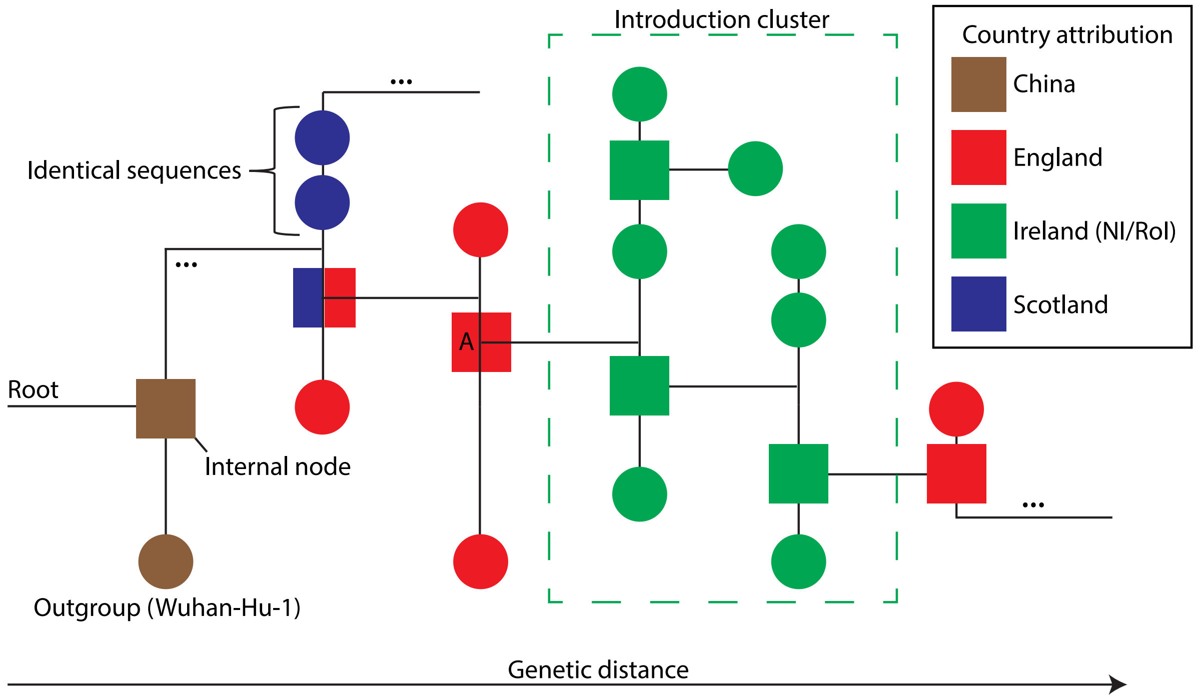


**Figure S3. Schematic phylogeny** An abstract representation of a phylogeny illustrating an Irish introduction cluster as analysed in the study. The ancestral node to the introduction cluster is emphasised with a red square (indicating inferred origin from England) and is marked with an 'A'. An internal node of ambiguous origin is depicted upstream of the ancestral node to the introduction cluster ('A'), to which it could not be discerned whether the internal node descends from Scotland or England (see blue and red square).

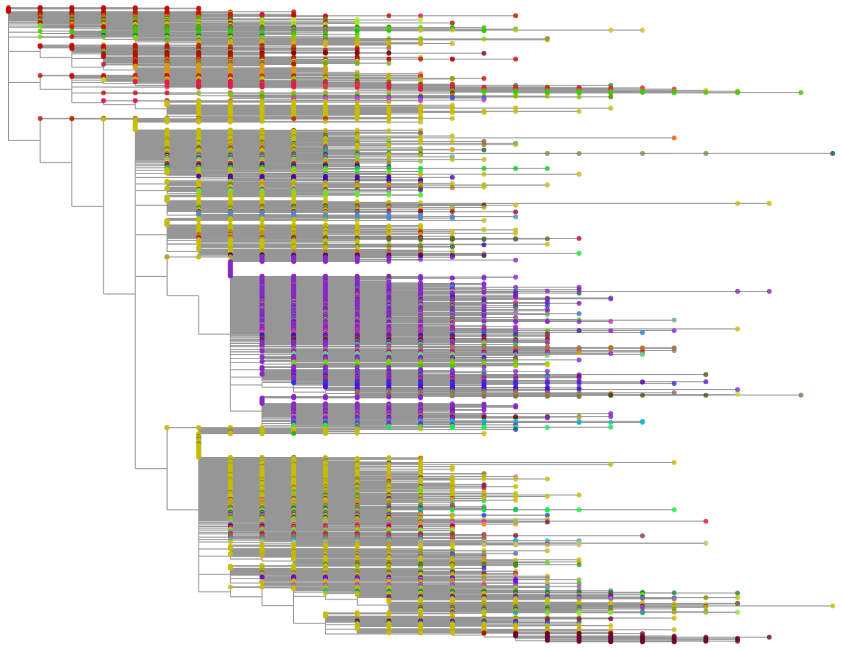

**Figure S4. Period A pruned tree**. Pruned SARS-CoV-2 phylogeny of 103,316 samples used for Period A analysis. Samples coloured by Pango (O'Toole et al. 2022) lineage and visualised using Taxonium (Sanderson 2022).

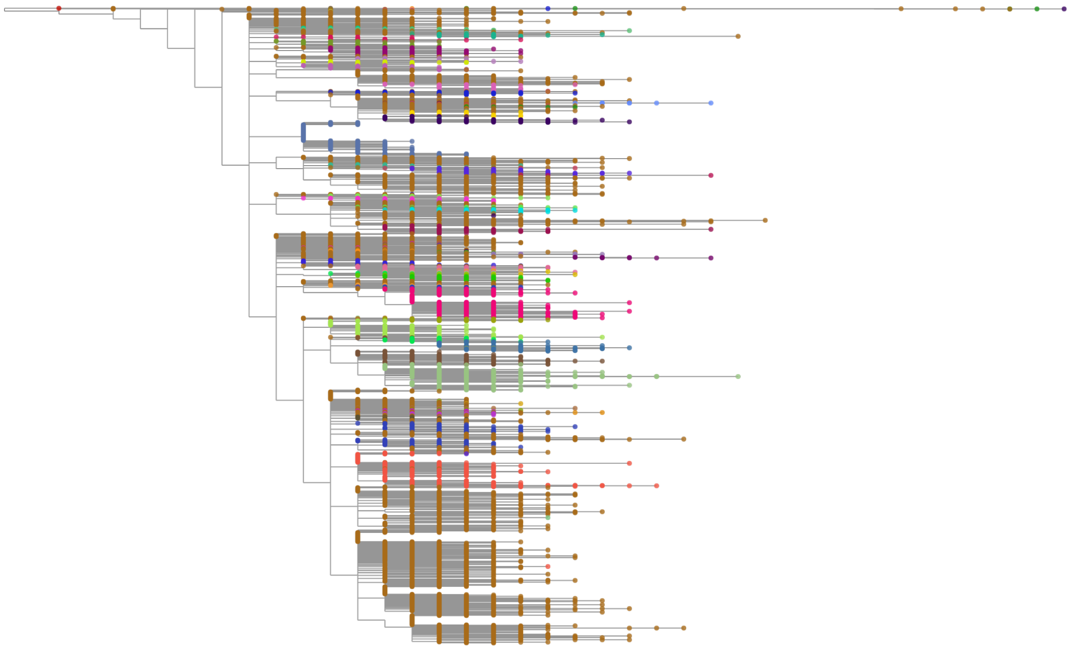

**Figure S5. Period B pruned tree**. Pruned SARS-CoV-2 phylogeny of 31,930 samples used for Period B analysis. Samples coloured by Pango (O'Toole et al. 2022) lineage and visualised using Taxonium (Sanderson 2022).

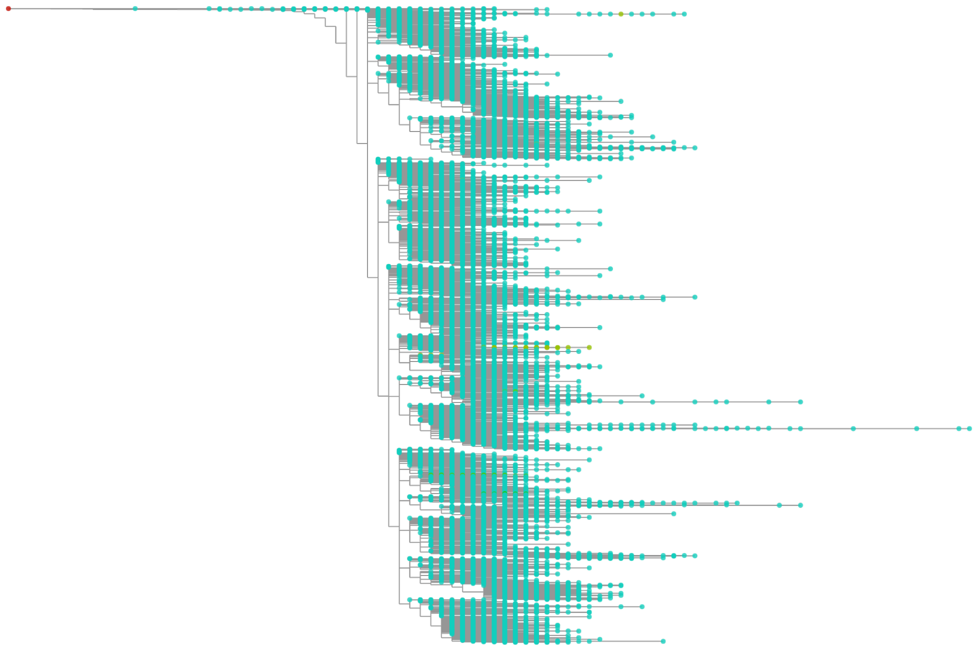

**Figure S6. Period C pruned tree**. Pruned SARS-CoV-2 phylogeny of 184,326 samples used for Period C analysis. Samples coloured by Pango (O'Toole et al. 2022) lineage and visualised using Taxonium (Sanderson 2022).

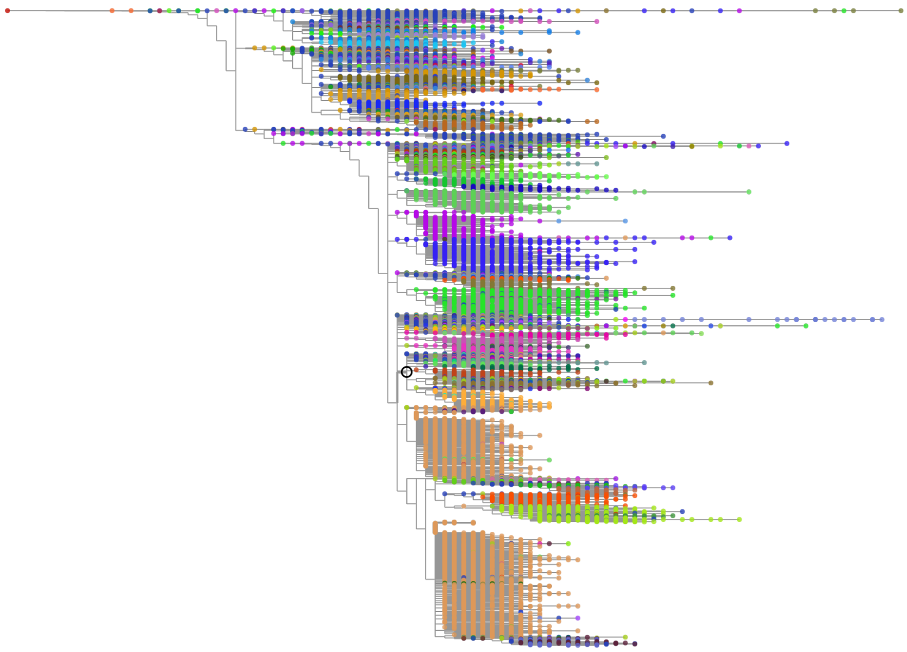

**Figure S7. Period D pruned tree**. Pruned SARS-CoV-2 phylogeny of 513,954 samples used for Period D analysis. Samples coloured by Pango (O'Toole et al. 2022) lineage and visualised using Taxonium (Sanderson 2022).

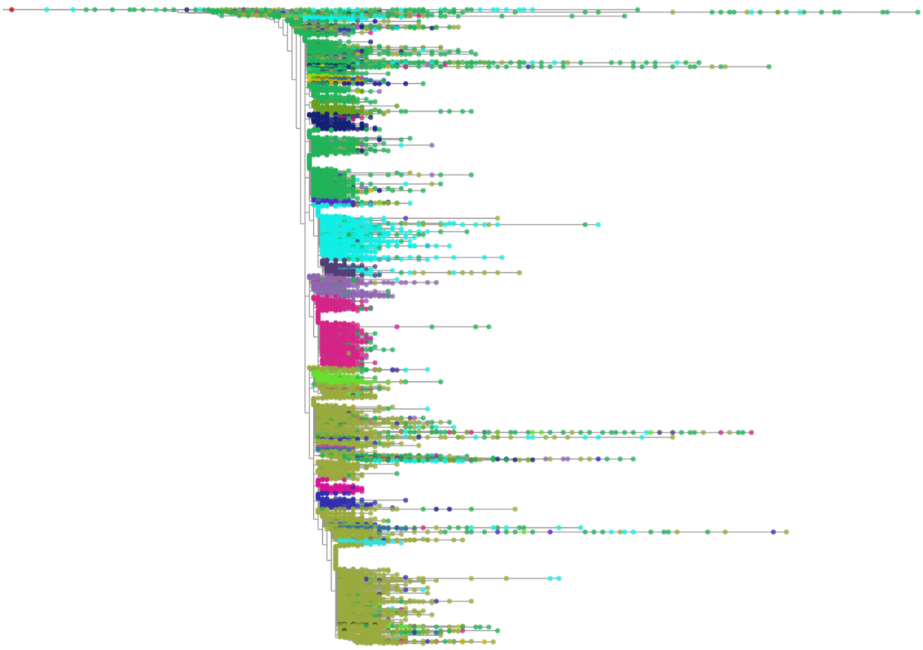

**Figure S8. Period E pruned tree**. Pruned SARS-CoV-2 phylogeny of 1,313,968 samples used for Period E analysis. Samples coloured by Pango (O'Toole et al. 2022) lineage and visualised using Taxonium (Sanderson 2022).

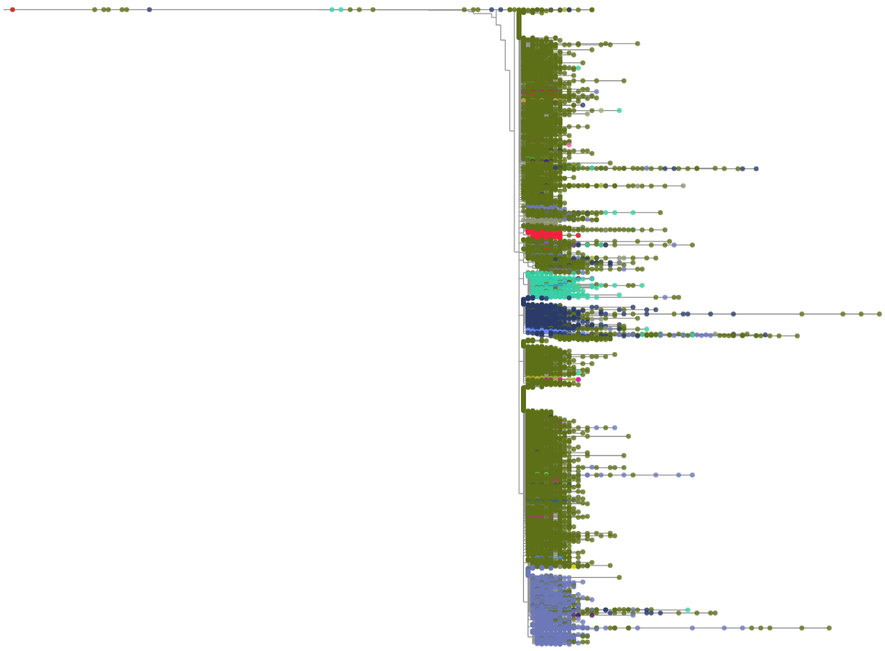

**Figure S9. Period F pruned tree**. Pruned SARS-CoV-2 phylogeny of 644,944 samples used for Period F analysis. Samples coloured by Pango (O'Toole et al. 2022) lineage and visualised using Taxonium (Sanderson 2022).

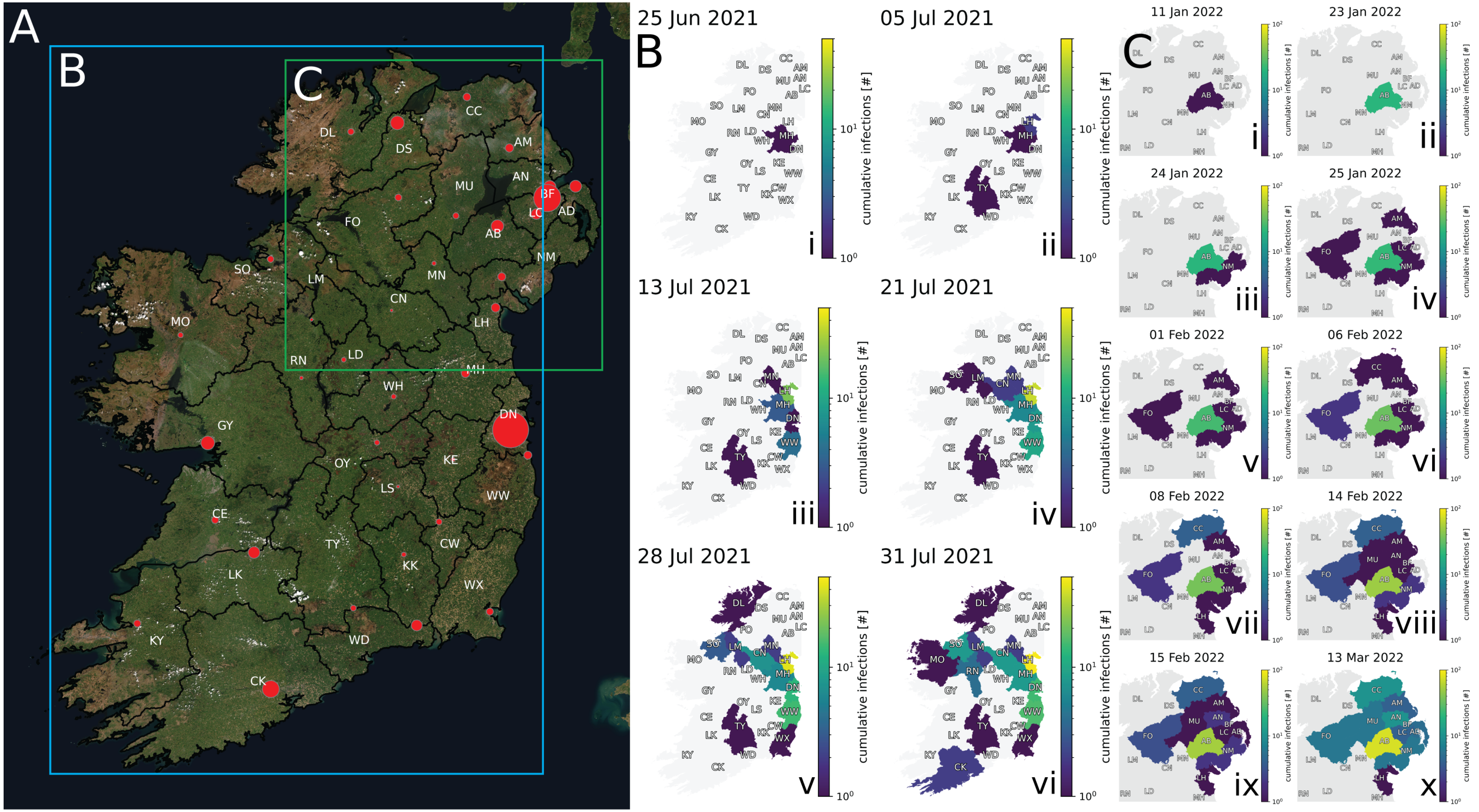


**Figure S10. Geospatial tracking of the spread of samples from an introduction event within Ireland A**, A map of Ireland created with GeoPandas v0.11.1 (Jordahl et al. (2022), `https://geopandas.org/`) using the Esri "World Imagery" basemap (Sources: Esri, DigitalGlobe, GeoEye, i-cubed, USDA FSA, USGS, AEX, Getmapping, Aerogrid, IGN, IGP, swisstopo, and the GIS User Community) as retrieved using contextily v1.2.0 (`https://github.com/geopandas/contextily`). Red points designate the locations of population centres in each government district with the size of each point scaled according to recent population estimates. Refer to Table S1 for the corresponding metadata for region abbreviations and population centres. Bounding boxes B (cyan) and C (green) depict the geographic extents of the two largest introduction clusters detected in this study, which are elucidated to the right. (caption continues on the following page.)

**Figure S10. Geospatial tracking of the spread of samples from an introduction event within Ireland (continued) B-i**, An introduction of Delta SARS-CoV-2 descended from England begins an introduction cluster in County Meath in RoI on 25th June 2021. **B-ii**, The introduction cluster exhibits a non-neighbouring spreading event into County Tipperary as well as adjacently to County Louth by 5th July 2021. **B-iii**, By 13th July 2021, the infection cluster has adjacently spread to County Monaghan, County Dublin, and County Waterford. **B-iv**, The introduction cluster reaches County Leitrim and County Sligo by 21st July 2021. **B-v**, County Donegal and County Wexford are reached by the introduction cluster by 28th July 2021. **B-vi**, The final geographic extent of the introduction cluster is observed, having reached County Cork, County Mayo, and County Roscommon by 31st July 2021. **C-i**, An introduction of BA.2 SARS-CoV-2 descended from Scotland is detected within the boundaries of Armagh City, Banbridge and Craigavon in NI on 11th January 2022. **C-ii**, 20 more infections afflict other individuals within the jurisdictional boundaries of Armagh City, Banbridge and Craigavon in NI through 23 January 2022. **C-iii**, An individual within the Newry, Mourne and Down is infected on 24th January 2022. **C-iv**, the infection cluster reaches three new NI local government districts, namely Fermanagh and Omagh, Mid and East Antrim, and Lisburn and Castlereagh on 25th January 2022. **C-v**, An affliction in Belfast belonging to the cluster is noted on 1st February 2022 **C-vi**, Spread of this importation cluster reaches Causeway Coast and Glens within on 6th February 2022. **C-vii**, The cluster reaches RoI by afflicting an individual in County Louth on 8th February 2022. **C-viii**, Further spreading occurs to Antrim and Newtownabbey and Mid Ulster **C-ix**, The clustered infection spread to its final geographic coverage having reached Ards and North Down on 15th February 2022 **C-x**, Spreading within the aforementioned regions continues until the last sequenced case in the cluster was recorded on 13th Mar 2022, indicating that this introduction cluster was tracked over 61 days.

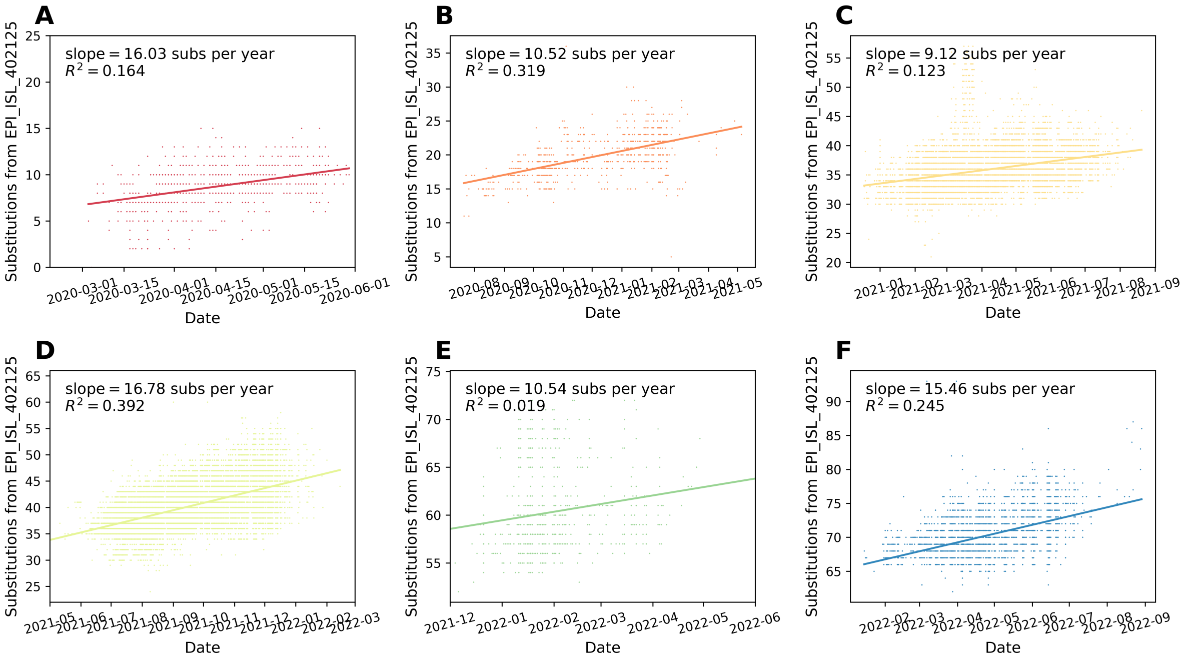


**Figure S11. OLS regressions of substitutions in Irish SARS-CoV-2 sequences over time per major introduction lineage**

Genome coverage for all samples included were at least 99.9% upon alignment to the Wuhan-Hu-1 SARS-CoV-2 reference (GISAID: EPI_ISL_402125). See Table S8 for related statistics. **A**, Initial introductions (Period A) **B**, B.1.177 (Period B) **C**, B.1.1.7 (Period C) **D**, Delta (Period D) **E**, Omicron (BA.1*) (Period E) **F**, Omicron (BA.2*) (Period F)

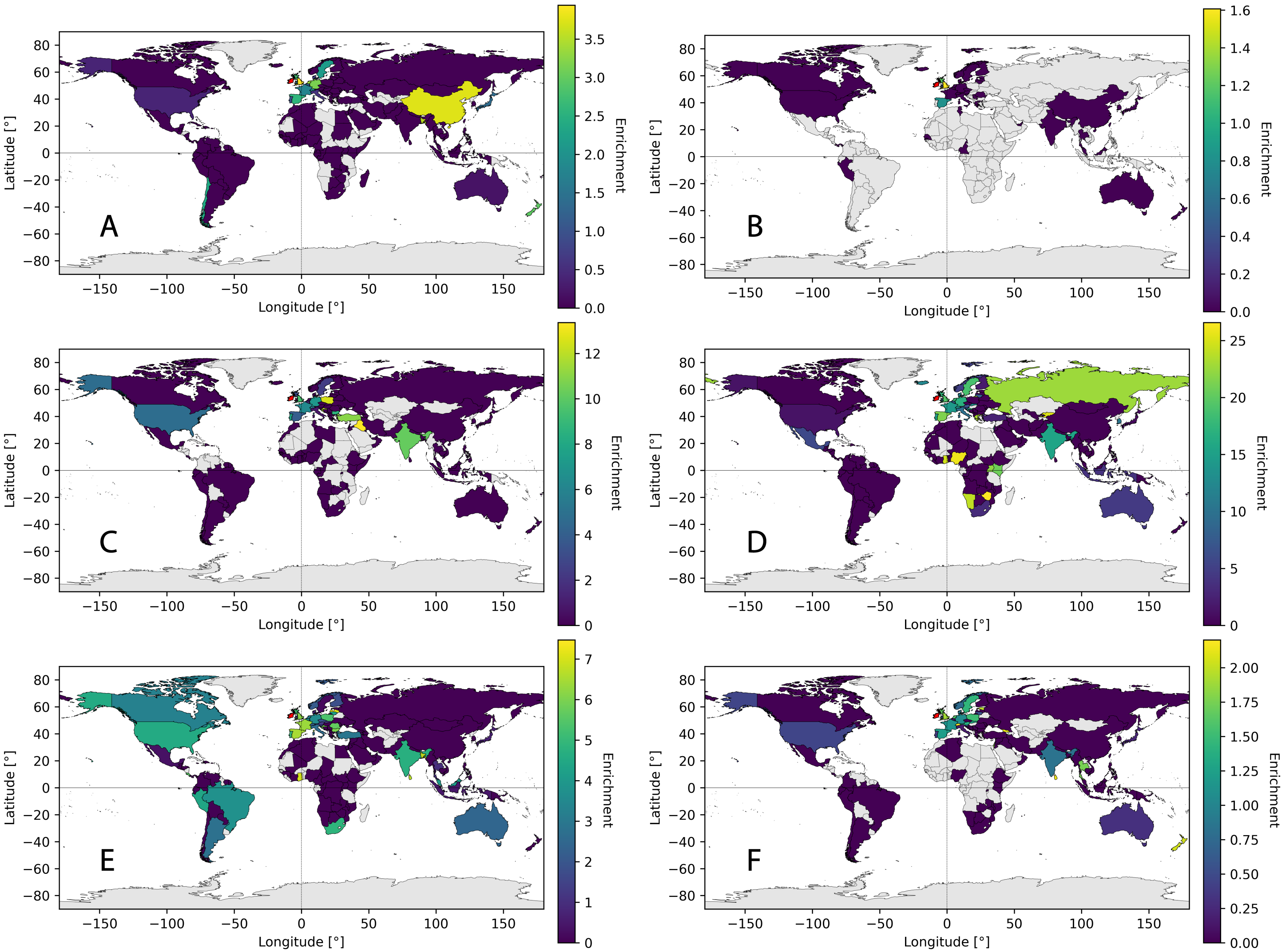


**Figure S12. World maps depicting the ratio of introductions to Ireland given the proportion of samples in the global phylogeny for each period** World maps were created by using and GeoPandas v0.11.1 (Jordahl et al. (2022) `https://geopandas.org/`). Sequence enrichment is defined as follows: Enrichment $= \frac{\text{proportion of introductions to Ireland}}{\text{proportion of tips in the global phylogeny}}$ (caption continues on the following page.)

**Figure S12. World maps depicting the ratio of introductions to Ireland given the proportion of samples in the global phylogeny for each period (continued)A**, A representation of global sequence enrichment from Period A. **B**, A representation of global sequence enrichment from Period B. **C**, A representation of global sequence enrichment from Period C. **D**, A representation of global sequence enrichment from Period D. **E**, A representation of global sequence enrichment from Period E. **F**, A representation of global sequence enrichment from Period F.

**Table S1.** Metadata for mapping samples to local government districts in Ireland (NI and RoI) Population data within NI and RoI were obtained from NISRA (`https://www.nisra.gov.uk/publications/census-2021-main-statistics-for-northern-ireland-phase-1`) and CSO (`https://data.cso.ie/table/FP001`), respectively. Troendle–Simpson–Skvortsov 2-letter abbreviation codes were devised for each local government district.

| Country | Local government district | Abbreviation | Area [km$^2$] | Population | Population centre (PC) | PC Population | PC Latitude | PC Longitude |
|---|---|---|---|---|---|---|---|---|
| NI | Antrim and Newtownabbey | AN | 571 | 145661 | Newtownabbey | 67200 | 54.6570°N | 5.9070°W |
| NI | Ards and North Down | AD | 457 | 163659 | Bangor | 63480 | 54.6600°N | 5.6700°W |
| NI | Armagh City, Banbridge and Craigavon | AB | 1347 | 218656 | Craigavon | 71560 | 54.4472°N | 6.3883°W |
| NI | Belfast | BF | 134 | 345418 | Belfast | 337210 | 54.5973°N | 5.9301°W |
| NI | Causeway Coast and Glens | CC | 1980 | 141746 | Coleraine | 25540 | 55.1320°N | 6.6685°W |
| NI | Derry City and Strabane | DS | 1237 | 150756 | Derry/Londonderry | 85080 | 54.9958°N | 7.3074°W |
| NI | Fermanagh and Omagh | FO | 2847 | 116812 | Omagh | 20000 | 54.6003°N | 7.2984°W |
| NI | Lisburn and Castlereagh | LC | 503 | 149106 | Lisburn | 49890 | 54.5120°N | 6.0310°W |
| NI | Mid and East Antrim | AM | 1046 | 138994 | Ballymena | 31270 | 54.8631°N | 6.2783°W |
| NI | Mid Ulster | MU | 1821 | 150293 | Dungannon | 15860 | 54.5029°N | 6.7696°W |
| NI | Newry, Mourne and Down | NM | 1619 | 182074 | Newry | 28410 | 54.1760°N | 6.3490°W |
| RoI | Carlow | CW | 895 | 61931 | Carlow | 16200 | 52.8360°N | 6.9245°W |
| RoI | Cavan | CN | 1856 | 81201 | Cavan | 3805 | 53.9943°N | 7.3608°W |
| RoI | Clare | CE | 3159 | 127419 | Ennis | 22100 | 52.8463°N | 8.9807°W |
| RoI | Cork | CK | 7255 | 581231 | Cork | 130119 | 51.8972°N | 8.4700°W |
| RoI | Donegal | DL | 4764 | 166321 | Letterkenny | 130119 | 54.9490°N | 7.7342°W |
| RoI | Dublin | DN | 924 | 1450701 | Dublin | 588233 | 53.3441°N | 6.2675°W |
| RoI | Galway | GY | 5796 | 276451 | Galway | 83456 | 53.2719°N | 9.0489°W |
| RoI | Kerry | KY | 4679 | 155528 | Tralee | 22300 | 52.2675°N | 9.6962°W |
| RoI | Kildare | KE | 1693 | 246977 | Newbridge | 7767 | 53.1805°N | 6.7959°W |
| RoI | Kilkenny | KK | 2061 | 103685 | Kilkenny | 9729 | 52.6537°N | 7.2480°W |
| RoI | Laois | LS | 1719 | 91657 | Portlaoise | 3920 | 53.0309°N | 7.3008°W |
| RoI | Leitrim | LM | 1502 | 35087 | Carrick-on-Shannon | 4062 | 53.9440°N | 8.0950°W |
| RoI | Limerick | LK | 2683 | 205444 | Limerick | 61570 | 52.6653°N | 8.6238°W |
| RoI | Longford | LD | 1040 | 46634 | Longford | 9600 | 53.7270°N | 7.7998°W |
| RoI | Louth | LH | 824 | 139100 | Dundalk | 35300 | 54.0090°N | 6.4049°W |
| RoI | Mayo | MO | 5351 | 137231 | Castlebar | 12068 | 53.8608°N | 9.2988°W |
| RoI | Meath | MH | 2332 | 220296 | Navan | 31800 | 53.6528°N | 6.6814°W |
| RoI | Monaghan | MN | 1273 | 64832 | Monaghan | 7678 | 54.2479°N | 6.9708°W |
| RoI | Offaly | OY | 1995 | 82668 | Tullamore | 11894 | 53.2739°N | 7.4945°W |
| RoI | Roscommon | RN | 2445 | 69995 | Roscommon | 5876 | 53.6279°N | 8.1886°W |
| RoI | Sligo | SO | 1791 | 69819 | Sligo | 18518 | 54.2706°N | 8.4716°W |
| RoI | Tipperary | TY | 4248 | 167661 | Clonmel | 14800 | 52.3539°N | 7.7116°W |
| RoI | Waterford | WD | 1836 | 127085 | Waterford | 54352 | 52.2567°N | 7.1292°W |
| RoI | Westmeath | WH | 1756 | 95840 | Mullingar | 10732 | 53.5258°N | 7.3412°W |
| RoI | Wexford | WX | 2353 | 163527 | Wexford | 22200 | 52.3342°N | 6.4575°W |
| RoI | Wicklow | WW | 2000 | 155485 | Bray | 28400 | 53.2044°N | 6.1092°W |

**Table S2.** Originating countries and their frequencies of importations for Period A. Rows in italics could not be fully resolved and are ambiguous.

| Origin country | Both | Into RoI | Into NI |
|---|---|---|---|
| Republic of Ireland (RoI) | – | – | 2 |
| Northern Ireland (NI) | – | 1 | – |
| England | 100 | 61 | 39 |
| USA | 17 | 12 | 5 |
| Scotland | 8 | 1 | 7 |
| Spain | 6 | 4 | 2 |
| France | 5 | 3 | 2 |
| Germany | 5 | 3 | 2 |
| Netherlands | 5 | 4 | 1 |
| China | 4 | 3 | 1 |
| Japan | 4 | 2 | 2 |
| Sweden | 2 | 1 | 1 |
| Switzerland | 2 | 1 | 1 |
| Wales | 2 | 1 | 1 |
| Portugal | 1 | 1 | 0 |
| Italy | 1 | 1 | 0 |
| Belgium | 1 | 1 | 0 |
| Chile | 1 | 0 | 1 |
| Bangladesh | 1 | 1 | 0 |
| Australia | 1 | 0 | 1 |
| New Zealand | 1 | 1 | 0 |
| Denmark | 1 | 1 | 0 |
| *Chile / RoI / England* | 1 | 1 | 0 |
| *Germany / Austria* | 1 | 0 | 1 |
| **Total into Ireland** | 170 | 103 | 67 |

**Table S3.** Originating countries and their frequencies of importations for Period B. Rows in italics could not be fully resolved and are ambiguous.

| Origin country | Both | Into RoI | Into NI |
|---|---|---|---|
| Republic of Ireland (RoI) | – | – | 0 |
| Northern Ireland (NI) | – | 0 | – |
| England | 34 | 13 | 21 |
| Scotland | 4 | 3 | 1 |
| Wales | 1 | 0 | 1 |
| Spain | 1 | 1 | 0 |
| *Latvia / Norway / Lithuania / Iceland* | 1 | 1 | 0 |
| **Total into Ireland** | 41 | 18 | 23 |

**Table S4.** Originating countries and their frequencies of importations for Period C. Rows in italics could not be fully resolved and are ambiguous.

| Origin country | Both | Into RoI | Into NI |
|---|---|---|---|
| Republic of Ireland (RoI) | – | – | 11 |
| Northern Ireland (NI) | – | 14 | – |
| England | 176 | 137 | 39 |
| Germany | 9 | 6 | 3 |
| USA | 5 | 5 | 0 |
| Poland | 4 | 3 | 1 |
| France | 4 | 4 | 0 |
| Sweden | 4 | 4 | 0 |
| Switzerland | 3 | 3 | 0 |
| Scotland | 3 | 2 | 1 |
| Spain | 2 | 2 | 0 |
| Turkey | 1 | 1 | 0 |
| Belgium | 1 | 1 | 0 |
| India | 1 | 1 | 0 |
| Croatia | 1 | 1 | 0 |
| Italy | 1 | 1 | 0 |
| Portugal | 1 | 1 | 0 |
| Iraq | 1 | 1 | 0 |
| *Germany / England* | 1 | 0 | 1 |
| **Total into Ireland** | 218 | 173 | 45 |

**Table S5.** Originating countries and their frequencies of importations for Period D. Rows in italics could not be fully resolved and are ambiguous.

| Origin country | Both | Into RoI | Into NI | *Into RoI/NI* |
|---|---|---|---|---|
| Republic of Ireland (RoI) | – | – | 42 | |
| Northern Ireland (NI) | – | 31 | – | |
| England | 473 | 244 | 228 | 1 |
| Scotland | 44 | 19 | 25 | |
| India | 38 | 33 | 5 | |
| Spain | 24 | 23 | 1 | |
| France | 15 | 15 | 0 | |
| Netherlands | 14 | 14 | 0 | |
| Denmark | 13 | 12 | 1 | |
| USA | 12 | 12 | 0 | |
| Wales | 12 | 4 | 8 | |
| Sweden | 11 | 11 | 0 | |
| Germany | 9 | 9 | 0 | |
| Russia | 7 | 6 | 1 | |
| Greece | 6 | 6 | 0 | |
| Italy | 6 | 5 | 1 | |
| Switzerland | 5 | 5 | 0 | |
| Belgium | 3 | 3 | 0 | |
| Portugal | 3 | 2 | 1 | |
| Nigeria | 3 | 3 | 0 | |
| Turkey | 2 | 2 | 0 | |
| Iceland | 2 | 2 | 0 | |
| Kenya | 1 | 1 | 0 | |
| Lithuania | 1 | 1 | 0 | |
| Hong Kong | 1 | 1 | 0 | |
| Australia | 1 | 1 | 0 | |
| Japan | 1 | 1 | 0 | |
| Uganda | 1 | 1 | 0 | |
| Namibia | 1 | 1 | 0 | |
| Wales—England | 1 | 0 | 1 | |
| Kyrgyzstan | 1 | 1 | 0 | |
| Croatia | 1 | 1 | 0 | |
| Israel | 1 | 1 | 0 | |
| Zimbabwe | 1 | 1 | 0 | |
| South Africa | 1 | 1 | 0 | |
| Slovakia | 1 | 1 | 0 | |
| Finland | 1 | 1 | 0 | |
| *England / RoI* | 4 | 4 | 0 | |
| *Italy / Belgium* | 1 | 1 | 0 | |
| *Netherlands / Denmark / RoI* | 1 | 1 | 0 | |
| *France / Belgium* | 1 | 1 | 0 | |
| *Italy / Denmark* | 1 | 1 | 0 | |
| *Germany / Israel* | 1 | 1 | 0 | |
| *Scotland / NI* | 1 | 0 | 1 | |
| *RoI / Portugal* | 1 | 1 | 0 | |
| **Total into Ireland** | 728 | 454 | 273 | 1 |

**Table S6.** Originating countries and their frequencies of importations for Period E. Rows in italics could not be fully resolved and are ambiguous.

| Origin country | Both | Into RoI | Into NI |
|---|---|---|---|
| Republic of Ireland (RoI) | – | – | 38 |
| Northern Ireland (NI) | – | 31 | – |
| England | 973 | 591 | 382 |
| USA | 513 | 379 | 134 |
| France | 121 | 105 | 16 |
| Scotland | 43 | 14 | 29 |
| Germany | 41 | 32 | 9 |
| Spain | 39 | 34 | 5 |
| Canada | 25 | 21 | 4 |
| Poland | 24 | 21 | 3 |
| Brazil | 13 | 10 | 3 |
| Wales | 12 | 7 | 5 |
| Croatia | 9 | 8 | 1 |
| India | 9 | 5 | 4 |
| Belgium | 8 | 8 | 0 |
| Denmark | 8 | 8 | 0 |
| Switzerland | 7 | 7 | 0 |
| Japan | 7 | 5 | 2 |
| Australia | 7 | 7 | 0 |
| South Africa | 7 | 4 | 3 |
| Netherlands | 6 | 6 | 0 |
| Italy | 6 | 4 | 2 |
| Portugal | 4 | 3 | 1 |
| Turkey | 3 | 3 | 0 |
| Romania | 3 | 2 | 1 |
| Slovenia | 3 | 2 | 1 |
| Latvia | 3 | 3 | 0 |
| Norway | 3 | 3 | 0 |
| Peru | 2 | 2 | 0 |
| Czech Republic | 2 | 2 | 0 |
| Lithuania | 2 | 2 | 0 |
| Bulgaria | 2 | 1 | 1 |
| Slovakia | 1 | 0 | 1 |
| Bangladesh | 1 | 1 | 0 |
| Sri Lanka | 1 | 1 | 0 |
| Mauritius | 1 | 1 | 0 |
| Ghana | 1 | 1 | 0 |
| Mexico | 1 | 1 | 0 |
| Finland | 1 | 1 | 0 |
| Sweden | 1 | 1 | 0 |
| Malaysia | 1 | 1 | 0 |
| Costa Rica | 1 | 1 | 0 |
| Seychelles | 1 | 1 | 0 |
| Argentina | 1 | 1 | 0 |
| Indonesia | 1 | 1 | 0 |
| Thailand | 1 | 1 | 0 |
| Austria | 1 | 1 | 0 |
| Reunion | 1 | 1 | 0 |
| *RoI / Poland* | 2 | 2 | 0 |
| *RoI / Germany* | 2 | 2 | 0 |
| *England / RoI* | 2 | 2 | 0 |
| *RoI / Switzerland* | 1 | 1 | 0 |
| *Netherlands / Poland* | 1 | 1 | 0 |
| *Scotland / NI* | 1 | 0 | 1 |
| *South Africa / Poland* | 1 | 1 | 0 |
| *RoI / Indonesia* | 1 | 1 | 0 |
| *France / Germany* | 1 | 1 | 0 |
| *RoI / Slovakia* | 1 | 1 | 0 |
| *Italy / Poland* | 1 | 1 | 0 |
| *Slovakia / Poland* | 1 | 1 | 0 |
| *RoI / USA* | 1 | 1 | 0 |
| **Total into Ireland** | 1,937 | 1,329 | 608 |

**Table S7.** Originating countries and their frequencies of importations for Period F. Rows in italics could not be fully resolved and are ambiguous.

| Origin country | Both | Into RoI | Into NI |
|---|---|---|---|
| Republic of Ireland (RoI) | – | – | 5 |
| Northern Ireland (NI) | – | 29 | – |
| England | 779 | 218 | 561 |
| Denmark | 137 | 41 | 96 |
| Scotland | 111 | 22 | 89 |
| Germany | 78 | 30 | 48 |
| India | 28 | 16 | 12 |
| Switzerland | 27 | 24 | 3 |
| Sweden | 26 | 9 | 17 |
| France | 18 | 9 | 9 |
| USA | 13 | 5 | 8 |
| Wales | 11 | 6 | 5 |
| Netherlands | 10 | 7 | 3 |
| Poland | 9 | 4 | 5 |
| Israel | 5 | 3 | 2 |
| Norway | 5 | 0 | 5 |
| Austria | 4 | 2 | 2 |
| Spain | 4 | 2 | 2 |
| Slovakia | 4 | 4 | 0 |
| Belgium | 2 | 2 | 0 |
| Thailand | 2 | 1 | 1 |
| Australia | 2 | 1 | 1 |
| Estonia | 2 | 0 | 2 |
| Hong Kong | 1 | 0 | 1 |
| Sri Lanka | 1 | 1 | 0 |
| Slovenia | 1 | 1 | 0 |
| Japan | 1 | 1 | 0 |
| Portugal | 1 | 1 | 0 |
| Georgia | 1 | 1 | 0 |
| New Zealand | 1 | 0 | 1 |
| Lithuania | 1 | 1 | 0 |
| Italy | 1 | 0 | 1 |
| *NI / Scotland* | 2 | 0 | 2 |
| *RoI / France* | 2 | 2 | 0 |
| *Germany / England* | 2 | 1 | 1 |
| *USA / England* | 1 | 1 | 0 |
| *NI / England* | 1 | 0 | 1 |
| *Scotland / RoI* | 1 | 1 | 0 |
| *Israel / France / Slovakia* | 1 | 1 | 0 |
| *USA / Wales* | 1 | 1 | 0 |
| *USA / NI* | 1 | 0 | 1 |
| *USA / RoI* | 1 | 1 | 0 |
| *Germany / NI* | 1 | 0 | 1 |
| *Netherlands / RoI / England* | 1 | 1 | 0 |
| *England / France* | 1 | 1 | 0 |
| *Poland / England* | 1 | 1 | 0 |
| *Germany/ RoI* | 1 | 1 | 0 |
| *Germany / France* | 1 | 0 | 1 |
| *France / Switzerland* | 1 | 1 | 0 |
| *England / Spain* | 1 | 1 | 0 |
| *RoI / England / Wales* | 1 | 1 | 0 |
| *RoI / England* | 1 | 1 | 0 |
| **Total into Ireland** | 1,309 | 428 | 881 |

**Table S8.** OLS linear regression statistics for estimating substitution rates of major imported lineages to Ireland.

| **NI** | **N** | **slope** | **standard error** | $R^2$ | $p$ |
|---|---|---|---|---|---|
| Period A (Initial Introductions) | 303 | 0.0438 | 0.0050 | 0.2047 | $1.078 \times 10^{-16}$ |
| Period B (B.1.177) | 285 | 0.0397 | 0.0022 | 0.5414 | $7.854 \times 10^{-50}$ |
| Period C (B.1.1.7) | 2409 | 0.0246 | 0.0012 | 0.1570 | $2.252 \times 10^{-91}$ |
| Period D (Delta) | 12025 | 0.0441 | 0.0006 | 0.3008 | ∼0 |
| Period E (Omicron (BA.1*)) | 186 | 0.0343 | 0.0045 | 0.2365 | $1.974 \times 10^{-12}$ |
| Period F (Omicron (BA.2*)) | 2172 | 0.0391 | 0.0019 | 0.1574 | $8.293 \times 10^{-83}$ |
| **RoI** | **N** | **slope** | **standard error** | $R^2$ | $p$ |
| Period A (Initial Introductions) | 476 | 0.0539 | 0.0056 | 0.1616 | $6.536 \times 10^{-20}$ |
| Period B (B.1.177) | 594 | 0.0229 | 0.0018 | 0.2066 | $1.292 \times 10^{-31}$ |
| Period C (B.1.1.7) | 8895 | 0.0254 | 0.0008 | 0.1139 | $6.896 \times 10^{-236}$ |
| Period D (Delta) | 17968 | 0.0465 | 0.0004 | 0.4096 | ∼0 |
| Period E (Omicron (BA.1*)) | 1347 | 0.0255 | 0.0070 | 0.0099 | $2.476 \times 10^{-4}$ |
| Period F (Omicron (BA.2*)) | 2612 | 0.0588 | 0.0018 | 0.2962 | $2.237 \times 10^{-201}$ |
| **Ireland (NI + RoI)** | **N** | **slope** | **standard error** | $R^2$ | $p$ |
| Period A (Initial Introductions) | 779 | 0.0439 | 0.0036 | 0.1637 | $4.803 \times 10^{-32}$ |
| Period B (B.1.177) | 879 | 0.0288 | 0.0014 | 0.3193 | $2.627 \times 10^{-75}$ |
| Period C (B.1.1.7) | 11304 | 0.0250 | 0.0006 | 0.1229 | ∼0 |
| Period D (Delta) | 29993 | 0.0460 | 0.0003 | 0.3922 | ∼0 |
| Period E (Omicron (BA.1*)) | 1533 | 0.028886 | 0.005237 | 0.0195 | $4.064 \times 10^{-8}$ |
| Period F (Omicron (BA.2*)) | 4784 | 0.0424 | 0.0011 | 0.2448 | $7.246 \times 10^{-294}$ |
